# Repeat Hospitalisation Following Admission for Mental Ill-health and Stress-Related Presentations in Children and Young People in England between 2014-2019: A Retrospective Cohort Study

**DOI:** 10.64898/2026.04.01.26349988

**Authors:** Caroline Skirrow, Matt Bird, Ellie Day, Jelena Savović, Frank de Vocht, Andy Judge, Paul Moran, Behnaz Schofield, Isobel Ward

**Affiliations:** Population Health Sciences, Bristol Medical School, University of Bristol, UK; The National Institute for Health and Care Research Applied Research Collaboration West (NIHR ARC West) at University Hospitals Bristol and Weston NHS Foundation Trust, UK; Emergency Medicine and Paediatric Emergency Medicine, North Bristol NHS Trust, UK; National Institute for Health and Care Research (NIHR) Bristol Evidence Synthesis Group, Population Health Sciences, University of Bristol, Bristol, UK; NIHR Biomedical Research Centre at University Hospitals Bristol and Weston NHS Foundation Trust and the University of Bristol, Bristol, UK; Centre for Academic Mental Health, Population Health Sciences Department, Bristol Medical School, University of Bristol, UK; University of the West of England (UWE), Bristol, UK; Nuffield Department of Orthopaedics, Rheumatology and Musculoskeletal Sciences, University of Oxford, Oxford, UK; Musculoskeletal Research Unit, Translational Health Sciences, Bristol Medical School, University of Bristol, Learning and Research Building, Level 1, Southmead Hospital, Bristol, BS10 5NB UK

**Keywords:** Mental health, hospital admissions, children and adolescent health, young adults, socio-economic deprivation

## Abstract

**Background:** Hospital admissions for mental health (MH) and stress-related presentations (SRP; symptoms without a clear medical cause which may be psychosomatic in nature) among children and young people (CYP) have risen over time. Re-hospitalisation contributes to service costs, may indicate gaps in community-based care, and can also disrupt education and social development.

**Methods:** This retrospective cohort study used NHS Hospital Episode Statistics to identify all CYP aged 10–25 with ≥1 MH/SRP-related hospital admissions in England between 1 April 2014 and 31 March 2018, with follow-up until 31 March 2019. Admissions were classified from ICD-10 codes into internalising, externalising, personality, and eating disorders, psychosis, self-harm, substance use, postpartum, or potentially psychosomatic diagnostic groups. Outcomes included 30-day all-cause readmission, 1-year all-cause readmission, and 1-year MH/SRP-specific re-hospitalisation. Time to re-hospitalisation, and number of MH/SRP readmissions were also evaluated. Clinical and sociodemographic characteristics associated with re-hospitalisation were assessed using regression models, time to re-hospitalisation using Kaplan-Meier analyses, and diagnostic transitions were visualised using Sankey diagrams.

**Results:** Of 492,061 CYP with hospital admission for MH/SRP, approximately one third were re-hospitalised within one year. Females, older CYP and those from more deprived areas had higher odds of all-cause readmission. The odds of MH/SRP rehospitalisation were highest among those aged 14–15 years. Co-occurring chronic physical health conditions, personality and eating disorders were associated with higher odds, and shorter time, to readmission.

**Conclusions:** Re-hospitalisation following MH/SRP admissions is common and socio-economically patterned among CYP. Targeted discharge planning and continuity of care interventions are needed, particularly for high-risk CYP admitted with eating and personality disorders.

## 1. Background

Admissions for mental health concerns among children and young people (CYP) have become increasingly prevalent, both in the UK (Ward et al., 2025) and in other high income countries (Soriano et al., 2025; Arakelyan et al. (2023), with sharp increases occurring during the pandemic (Overhage et al., 2023; Guitierrez-Sacristan et al., 2022). Repeated admissions are of clinical and policy relevance, because they may signal gaps in clinical or community care, while also placing additional pressures on health services. For CYP, repeated hospitalisation can disrupt education, employment and social development during critical life stages.

Prior systematic reviews have identified elevated risks of re-hospitalisation among CYP with a history of suicidal ideation, psychotic disorders, bipolar and mood disorders, developmental disorders (including autism and intellectual disability), eating disorders and personality disorders (Edgcomb, Sorter, Lorberg, and Zima, 2020). In contrast, CYP with substance use disorders have historically experienced lower re-hospitalisation risk (Edgcomb et al., 2020). Associations with demographic characteristics such as age, ethnicity and family income are less consistent across studies (Edgcomb et al., 2020; Madden et al., 2020).

Beyond clinical and demographic factors, a range of contextual and system-level influences have been identified as potentially contributing to re-hospitalisation. These include regional and environmental factors (e.g. deprivation and urbanicity), healthcare system characteristics (e.g. organisation and financing), hospital characteristics (e.g. capacity) and aspects of care provision such as post-discharge follow-up and care (Kalseth, Lassemo, Wahlbeck, Haaramo and Magnussen, 2016). As a result, re-hospitalisation is sometimes used as an indicator of care quality, given that repeated admissions may be considered potentially avoidable with effective treatment, discharge planning and continuity of care (Donisi, Tedeschi, Wahlbeck, Haaramo and Amaddeo, 2016; Sfetcu et al., 2017).

Recently, research focusing on CYP has adopted a broader conceptualisation of stress-related presentations (SRPs), encompassing not only mental health, substance use, and self-harm, but also unexplained pain, functional symptoms and other presentations without a clear medical cause which may be psychosomatic in nature (Blackburn et al., 2021; Geist, Weinstein, Walker, and Campo, 2008). Using this broader definition, studies suggest that in England, between 2014–2020, nearly one-third of emergency hospital admissions among 11–17 year-olds were related to mental health or SRPs (Ni Chobhthaigh, Jay and Blackburn, 2025), with around one-in-four between 2017-18 having multiple admissions in one year (Blackburn et al., 2021).

This demonstrates a significant burden from mental health and stress related presentations on both healthcare systems and the affected young people and their families. Yet, to date, there has been no detailed national investigation identifying factors associated with re-hospitalisation among CYP following presentation with mental health or stress related admissions. This study aims to address this gap: examining patterns, timing, sociodemographic and clinical factors associated with re-hospitalisation among CYP aged 10-25 in England.

## 2. Methods

### 2.1 Study design and population

This study included CYP (from age 10 until their 24th birthday) with at least one NHS hospital admission for mental health or stress-related presentations (MH/SRP) between 1st April 2014 and 31st March 2018. CYP were followed-up until 31 March 2019 up to the month of their 25^th^ birthday.

Hospital admissions were identified from Hospital Episode Statistics Admitted Patient Care (HES APC) database, an administrative dataset collected for the purposes of reimbursement, management and planning of services. HES APC collects information from all emergency and planned admissions to NHS hospitals which require a hospital bed across England, as well as for independent sector providers where costs are met by the NHS (Herbert, Wijlaars, Zylbersztejn, Cromwell, and Hardelid, 2017). HES APC does not capture attendance at accident and emergency departments or outpatient bookings, which are held separately.

Submission of data from mental health hospitals became less consistent and complete in HES APC since 2019 when reimbursement became contingent on data submission via the Mental health Services Dataset (Villasenor et al., 2023). The current study therefore focused on initial admissions occurring between 1st of April 2014 and 31st March 2018, capturing re-hospitalisations over this period and until 31st of March 2019.

### 2.2 Procedures

#### 2.2.1 Hospital admissions for Mental Health (MH) and Stress-Related Presentations (SRP)

HES-APC records are composed of hospital episodes of care under a single consultant within a hospital provider. Consistent with previous work, new episodes within one day of the end of the previous episode were considered part of the same inpatient spell (Herbert, Gilbert, Gonzalez-Izquierdo and Li, 2015; Ni Chobhthaigh et al., 2025). Discharge date was defined as the last completion date of any episode within a hospital spell. For each multi-episode spell, diagnosis on admission was identified based on International Classification of Diseases version 10 (ICD-10) codes from the first completed episode. However, if diagnostic information was missing or not specified (e.g. code R69), diagnosis was identified from subsequent episodes (Bottle et al., 2010).

Admissions due to mental health concerns or stress related presentations (MH/SRP presentations) were identified from ICD-10 codes, and grouped into externalising disorders, internalising disorders, psychosis, self-harm, eating disorders, substance use, postpartum, personality disorder, or potentially psychosomatic presentations. These groups were identified following established code lists and procedures (Ni Chobhthaigh et al., 2025), updated to include presentations for the older age-range in the current study in consultation with a consultant psychiatrist (author PM).

ICD-10 codes for MH/SRP presentations were required in the primary diagnosis field in HES APC entries, except for self-harm, where codes could appear in any diagnostic position (Ni Chobhthaigh et al., 2025). This exception reflects prior research consistently showing that self-harm is rarely recorded as the primary reason for admission in HES (Ward et al., 2025). Admissions were only categorised as potentially psychosomatic where no identifiable medical, surgical or pregnancy-related causes for presentation was indicated within the same admission, as defined using established code lists (Harron, Gilbert, Cromwell and van der Meulen, 2016; Ni Chobhthaigh et al., 2025). Code lists are provided in Appendix S1 (Tables S1.1-S1.5) and [GitHub](https://github.com/ARCWest-ADS/HES_HIU.git).

#### 2.2.2 Index visits, and re-hospitalisations

The first index visit for each patient was defined as their first MH/SRP admission during the study period, from age 10 to up to the month of their 24th birthday. Multiple index admissions per patient were permitted to enable the identification of age-based trends in readmission patterns, provided that each new index visit occurred more than 365 days after a discharge from any previous spell.

Only index visits in which the patient survived to discharge were included. Follow-up was censored at end of March 2019 or the month of the patients’ 25th birthday.

Re-hospitalisations were defined as any new hospital spell starting 2–365 days after discharge from an index spell. An overview of data cleaning and sample flow is provided in Appendix S1, Figure SF1.

### 2.3 Outcome measures

#### 2.3.1 Binary re-hospitalisation indicators

Three primary outcome measures were evaluated to capture patterns of re-hospitalisation following index admissions:

1. All cause 30-day rehospitalisation: Readmission for any reason within 30 days of discharge following index admission.
2. All-cause 1-year rehospitalisation: Readmission for any reason within 365 days of discharge following index admission.
3. MH/SRP related 1-year rehospitalisation: Readmission for MH/SRP within 365 days of discharge following index admission.

#### 2.3.2 Time to re-hospitalisation

Time to re-hospitalisation was defined as the number of months between discharge from an index visit to the next readmission for any reason within one year.

#### 2.3.3 Number of readmissions

For individuals who experienced one or more re-hospitalisations within one year of discharge from index admission, the total number of re-hospitalisations for MH/SRP presentations within 365 days of each index admission was recorded.

### 2.4 Covariates

Age at each index visit was categorised into age bands (10-11, 12-13, 14-15, 16-17, 18-19, 20-21, 22-24 years). Ethnic group was categorised as White, Black, Asian, Mixed, or Other (Shiekh et al., 2023), taking the modal category up to and including the index spell; if this was non-specific (e.g., ‘Other’), the next most frequent category was used. Where no clear group was present, then the category at the index visit, or if missing, the most recent prior specific entry, was carried forward. Area-level deprivation was defined using population weighted index of multiple deprivation (IMD) quintile. Geographical region was derived from patients’ postcodes using Government Office Regions. Both measures were obtained from data recorded at the index visit; where missing, the closest prior available value was used.

A binary indicator for chronic physical health conditions in any diagnostic position, for any episodes up to and inclusive of index visits, was derived using ICD-10 code list developed by Hardelid, Dattani and Gilbert (2014), after excluding codes for mental health and substance use (Appendix S1: code lists, Table ST1.6). Discharge method (formal discharge vs. self-discharge) and admission method (emergency vs. non-emergency admission) were also defined from index admission. Missing data is summarised in ST1.

### 2.5 Statistical analysis

#### 2.5.1 Descriptive statistics

Counts and proportions of re-hospitalisations were calculated per index visit, and stratified by age, sex, ethnicity, region and IMD quintile, and by admission and discharge method and year.

#### 2.5.2 Diagnosis at index and re-hospitalisation

Sankey diagrams were used to visualise changes in diagnostic group between index visits and subsequent re-hospitalisations.

#### 2.5.3 Predicting re-hospitalisation: Generalised Estimating Equations

Clinical and demographic characteristics associated with re-hospitalisation were evaluated using Generalised Estimating Equations (GEE) with a logit link. Predictors of interest included diagnostic group, chronic physical health condition, admission and discharge method, and age, sex, ethnicity, IMD and region.

Analyses were carried out on complete data with defined categories and using an exchangeable correlation structure and robust (sandwich) standard errors to obtain 95% confidence intervals.

Models were run including the covariate of interest and fiscal year (minimally adjusted model), and accounting for all sociodemographic and clinical factors of interest (fully adjusted model). Odds ratios and 95% confidence intervals (95% CI) were derived.

#### 2.5.4 Time to readmission

Cumulative probability of readmission was estimated from Kaplan-Meier curves for each diagnostic group and logrank tests were used to compare differences between groups. Sensitivity analyses for Kaplan-Meier estimates were conducted selecting the first index visit only and excluding repeated index visits.

#### 2.5.5 Frequency of MH/SRP rehospitalisation

Restricting our cohort to just those with at least one MH/SHP readmission, Poisson regression models were used to estimate the clinical and demographic factors associated with higher rates of readmission, using the same covariates as in the GEE models. Readmission counts were capped at the 99th centile to limit the influence of extreme outliers. Analyses were carried out on complete data with defined covariate categories, as described above. Incident Rate Ratios (IRRs), alongside robust (sandwich) 95% confidence intervals were derived.

All data cleaning, preparation and analyses were completed with R software for Windows, version 4.4.3 (R Foundation for Statistical Computing, Vienna, Austria; see https://www.r-project.org/).

### 2.6 Ethics statement

De-identified NHS Hospital Episode Statistics data were provided to the researchers by NHS England within the terms of a data-sharing agreement (DARS-NIC-17875-X7K1V-v5.2). Ethical review and approval for the study and use of de-identified administrative data was provided by the Research Ethics Review Committee at the University of Bristol (reference: 27364).

## 3. Results

### 3.1 Descriptive Statistics

492,061 CYP with at least one qualifying MH/SRP index presentation were identified from records. MH/SRP index presentations in those aged 10-24 made up 27% of all-cause index presentations documented in HES-APC over the same time period (N=518,504 MH/SRP index presentations out of N=1,919,820 all-cause index admissions in CYP aged 18-24 between 2014-2018): defined as the number of patients with an admission for any reason, together with any subsequent admissions occurring at least 365 days after their most recent spell discharge.

The majority of MH/SRP index admissions were in females (64.97%) and 83.83% (n=434,674) of MH/SRP index presentations were emergency admissions. The most prevalent presentation category was ‘potentially psychosomatic’ (67.8%), followed by self-harm (25.4%; Table 1). Most patients were formally discharged following an index admission (n=502,938, 97.1%), and only few self-discharged or were discharged by another person (e.g. family member) outside of the health or legal system (n=14,148, 2.7%). All-cause rehospitalisation within 1-year of a MH/SRP index admission was observed in 36.1% (n=177,516) of CYP.

**Table 1:** Number and percentage of patients with index admissions and re-hospitalisations (Number (N) and percentage (%)), within 30 days, 1 year all-cause, and within 1 year for reasons associated with MH/SRP. IMD: Index of Multiple Deprivation, MH/SRP: Mental Health/Stress-related presentations. † Reported on a per-index event basis rather than a per-patient basis, because some patients have multiple index spells over time. * Counts suppressed for disclosure control.

| Group |  | Total population at index admission |  | 30-day all-cause re-hospitalisation |  | 1-year all-cause re-hospitalisation |  | 1-year SRP/MH only re-hospitalisation |  |
| --- | --- | --- | --- | --- | --- | --- | --- | --- | --- |
|  |  | N | % of total | N | % of index | N | % of index | N | % of index |
| **Sex** | Female | 319,716 | 65.0 | 35,846 | 11.2 | 128,544 | 40.2 | 69,037 | 21.6 |
|  | Male | 172,345 | 35.0 | 15,506 | 9.0 | 48,972 | 28.4 | 25,685 | 14.9 |
| **Ethnicity** | Asian | 30,377 | 6.8 | 3,563 | 11.7 | 10,729 | 35.3 | 5,188 | 17.1 |
|  | Black | 16,063 | 3.6 | 1,867 | 11.6 | 6,164 | 38.4 | 3,090 | 19.2 |
|  | Mixed | 10,643 | 2.4 | 1,154 | 10.8 | 4,055 | 38.1 | 2,234 | 21.0 |
|  | Other | 12,484 | 2.8 | 1,168 | 9.4 | 3,826 | 30.7 | 1,917 | 15.4 |
|  | White | 374,582 | 84.3 | 41,079 | 11.0 | 144,502 | 38.6 | 77,810 | 20.8 |
| **IMD** | 1 - most deprived | 137,009 | 28.3 | 15,301 | 11.2 | 54,976 | 40.1 | 28,700 | 21.0 |
|  | 2 | 106,963 | 22.1 | 11,192 | 10.5 | 39,802 | 37.2 | 20,957 | 19.6 |
|  | 3 | 88,987 | 18.4 | 9,242 | 10.4 | 31,665 | 35.6 | 17,036 | 19.1 |
|  | 4 | 77,525 | 16.0 | 7,907 | 10.2 | 26,397 | 34.1 | 14,275 | 18.4 |
|  | 5 - least deprived | 74,199 | 15.3 | 7,403 | 10.0 | 23,788 | 32.1 | 13,269 | 17.9 |
| **Region** | East Midlands | 36,013 | 7.3 | 3,719 | 10.3 | 13,619 | 37.8 | 7,294 | 20.3 |
|  | East of England | 48,174 | 9.8 | 5,029 | 10.4 | 17,387 | 36.1 | 9,355 | 19.4 |
|  | London | 69,263 | 14.1 | 7,093 | 10.2 | 24,088 | 34.8 | 12,342 | 17.8 |
|  | North East | 27,151 | 5.5 | 2,707 | 10.0 | 10,049 | 37.0 | 5,060 | 18.6 |
|  | North West | 79,435 | 16.1 | 8,632 | 10.9 | 30,028 | 37.8 | 15,930 | 20.1 |
|  | South East | 75,047 | 15.3 | 7,498 | 10.0 | 25,935 | 34.6 | 14,317 | 19.1 |
|  | South West | 50,409 | 10.2 | 5,387 | 10.7 | 18,314 | 36.3 | 10,620 | 21.1 |
|  | West Midlands | 57,607 | 11.7 | 5,980 | 10.4 | 20,148 | 35.0 | 10,832 | 18.8 |
|  | Yorkshire and Humber | 48,962 | 10.0 | 5,307 | 10.8 | 17,948 | 36.7 | 8,972 | 18.3 |
| **Age †** | 10-11 | 37,759 | 7.3 | 3,902 | 10.3 | 10,490 | 27.8 | 5,057 | 13.4 |
|  | 12-13 | 47,224 | 9.1 | 4,826 | 10.2 | 14,609 | 30.9 | 8,793 | 18.6 |
|  | 14-15 | 73,665 | 14.2 | 7,251 | 9.8 | 24,635 | 33.4 | 16,707 | 22.7 |
|  | 16-17 | 74,939 | 14.5 | 6,989 | 9.3 | 25,336 | 33.8 | 14,831 | 19.8 |
|  | 18-19 | 87,266 | 16.8 | 8,603 | 9.9 | 31,549 | 36.2 | 16,608 | 19.0 |
|  | 20-21 | 96,583 | 18.6 | 9,813 | 10.2 | 36,377 | 37.7 | 17,383 | 18.0 |
|  | 22-24 | 101,068 | 19.5 | 10,421 | 10.3 | 39,476 | 39.1 | 17,960 | 17.8 |
| **Index diagnosis group †** | Eating disorders | 3,357 | 0.7 | 610 | 18.2 | 1,506 | 44.9 | 1,257 | 37.4 |
|  | Externalising | 560 | 0.1 | * | * | 170 | 30.4 | * | * |
|  | Internalising | 8,472 | 1.6 | 768 | 9.1 | 2,783 | 32.9 | 1,730 | 20.4 |
|  | Personality disorders | 1,019 | 0.2 | 172 | 16.9 | 524 | 51.4 | 431 | 42.3 |
|  | Postpartum mental health | 75 | 0.01 | * | * | 16 | 21.3 | * | * |
|  | Potentially psychosomatic | 351,759 | 67.8 | 36,584 | 10.4 | 123,577 | 35.1 | 53,560 | 15.2 |
|  | Psychosis | 6,905 | 1.3 | 722 | 10.5 | 2,581 | 37.4 | 1,983 | 28.7 |
|  | Self-harm | 131,653 | 25.4 | 12,296 | 9.3 | 48,254 | 36.7 | 36,543 | 27.8 |
|  | Substance use and abuse | 14,704 | 2.8 | 607 | 4.1 | 3,061 | 20.8 | 1,718 | 11.7 |
| **chronic physical health condition †** | Not present at index | 336,197 | 64.8 | 22,197 | 6.6 | 81,904 | 24.4 | 49,317 | 14.7 |
|  | Present at index | 182,307 | 35.2 | 29,608 | 16.2 | 100,568 | 55.2 | 48,022 | 26.3 |
| **Admission type †** | Emergency | 434,674 | 83.9 | 43,246 | 10.0 | 149,898 | 34.5 | 82,528 | 19.0 |
|  | Non-emergency | 83,252 | 16.1 | 8,505 | 10.2 | 32,353 | 38.9 | 14,655 | 17.6 |
| **Discharge type †** | Discharged | 502,938 | 97.3 | 50,163 | 10.0 | 176,683 | 35.1 | 94,101 | 18.7 |
|  | Self-discharged | 14,148 | 2.7 | 1,477 | 10.4 | 5,259 | 37.2 | 2,866 | 20.3 |
| **Fiscal Year of index visit †** | 2014-2015 | 142,816 | 27.5 | 15,502 | 10.9 | 54,614 | 38.2 | 29,923 | 21.0 |
|  | 2015-2016 | 128,096 | 24.7 | 12,537 | 9.8 | 44,468 | 34.7 | 23,127 | 18.1 |
|  | 2016-2017 | 123,051 | 23.7 | 11,715 | 9.5 | 41,884 | 34.0 | 21,806 | 17.7 |
|  | 2017-2018 | 124,541 | 24.0 | 12,051 | 9.7 | 41,506 | 33.3 | 22,483 | 18.1 |

Within one year of index admission, the number of all-cause re-hospitalisations per individual ranged from 0 to over 130; 19.5% of index visits were followed by a single re-hospitalisation and 15.9% by two or more. When restricted to MH/SRP-related readmissions, counts ranged from 0 to over 30.

Overall, 19.0% of index admissions were followed by at least one MH/SRP re-hospitalisation, and 6.4% were followed by two or more such re-hospitalisations.

Around a third of patients with an MH/SRP index presentation had a co-occurring or pre-existing chronic physical health condition (35.5%), with the three most common being asthma or chronic lower respiratory diseases, digestive conditions, and renal or genito-urinary conditions. CYP with index presentations noted as potentially psychosomatic and those with postpartum mental health related conditions had the highest incidence of pre-existing or co-occurring chronic physical health conditions (Appendix S2, Table ST2).

### 3.2 Diagnosis at index and re-hospitalisation

47.2% of readmissions within 1-year were for the same diagnostic grouping recorded at time of index presentation (Appendix S1, tables ST3). Index admissions for potentially psychosomatic presentations were most frequently followed by re-hospitalisation unrelated to MH/SRP (Figure 1, Appendix S1 Figures SF2-SF3).

**Figure 1:**
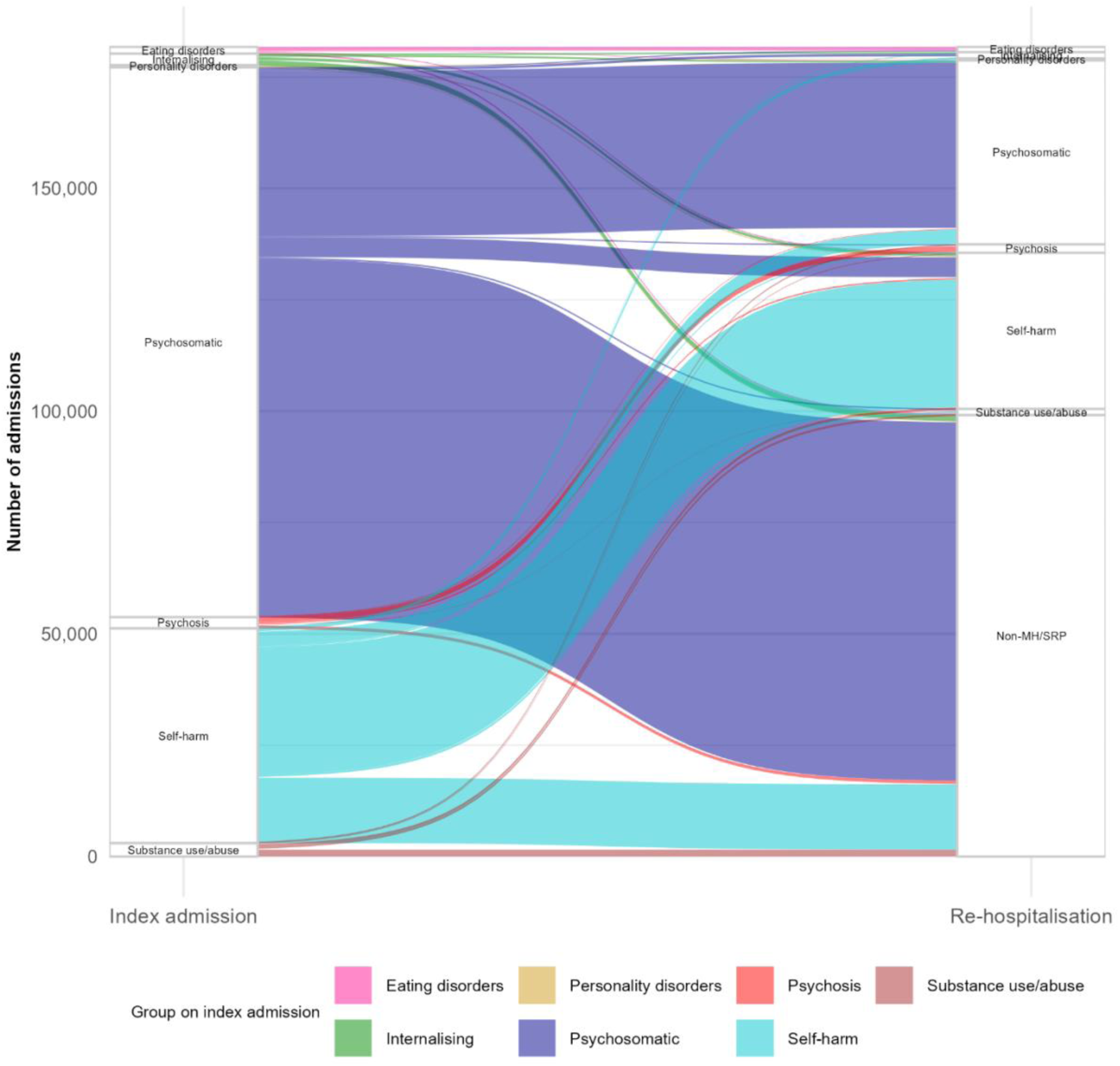
Alluvial plot showing transition and stability of the diagnostic groupings on successive admissions from index to all-cause re-hospitalisation within one year*. * with minimum transition frequency reported ≥100

The highest consistency of diagnostic grouping between index and successive admissions was observed for self-harm (59.4%), with rates comparatively high for eating disorders (52.7%) and psychosis (47.6%, Appendix S2 Table ST4).

### 3.3 Time to readmission

Cumulative incidence plots showed that re-hospitalisation risk was highest within the first month after discharge, accounting for around a third of all readmissions, and around half occurring within three months (Figure 2). Twelve-month cumulative probability or readmission was highest for personality disorders (51.4%, 95%CI 48.3-54.4) and eating disorders (44.9%, 95%CI 43.2-46.5) and lowest for substance use disorders (20.8%, 95%CI 20.2-21.5). Group differences were significant (log rank: Χ^2^=1833, p< 0.001), and *post-hoc* pairwise tests indicated higher readmission risk for personality disorders and lower risk for externalising disorders relative to the other groups (p<0.001, Bonferroni corrected). Findings were unchanged after removing repeated observations within individuals (Appendix S1 Figure SF3).

**Figure 2:**
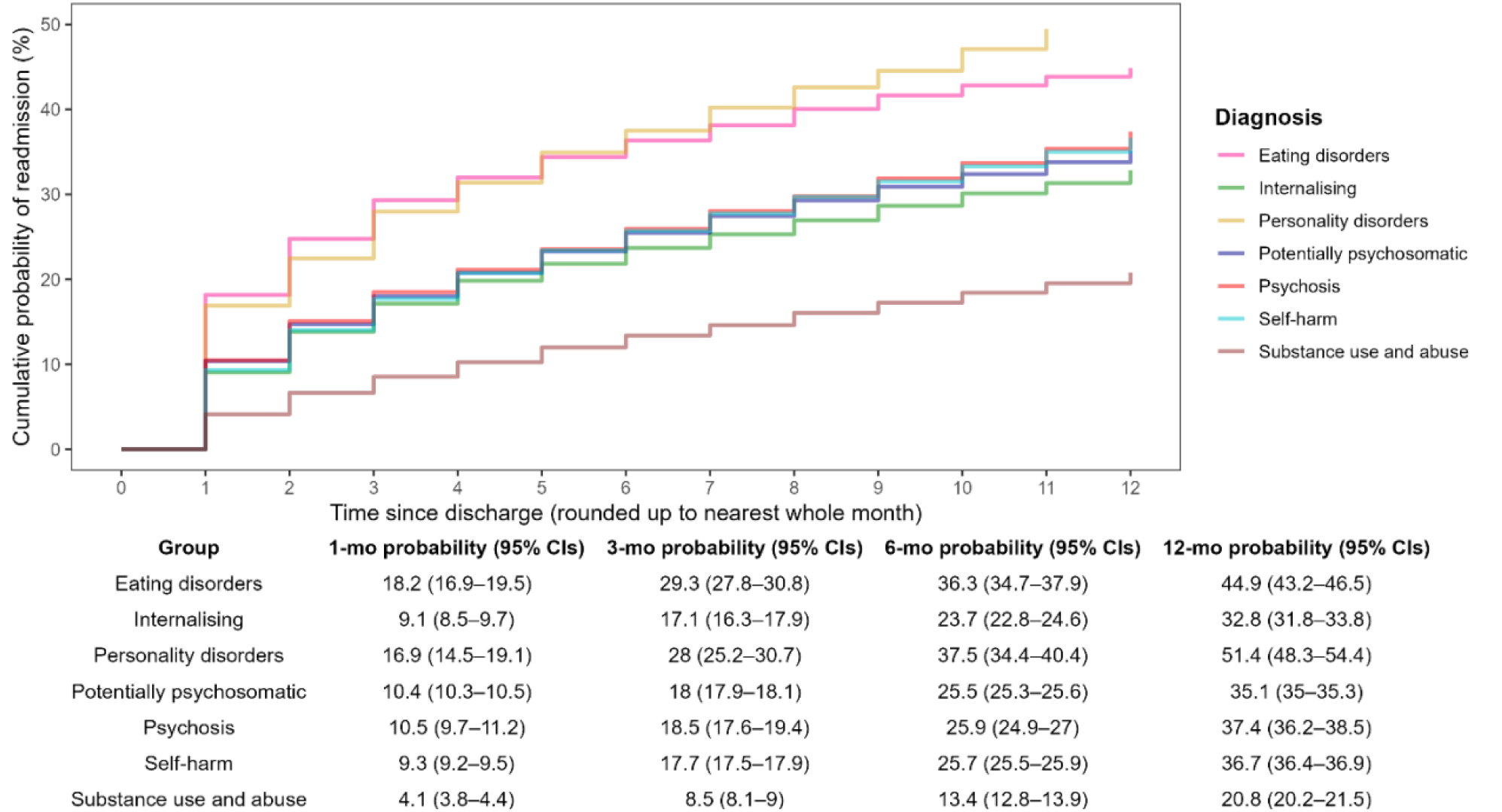
Cumulative monthly probability of all-cause readmission. Probability of diagnostic group stratified by diagnostic group with tabulation below showing 1-month, 3-month and 12-month readmission probability, together with 95% confidence intervals. Externalising and Postpartum MH groups suppressed to avoid disclosure.

### 3.4 Predictors of binary re-hospitalisation indicators

Being female was associated with significantly higher odds of readmission across all outcomes (30-day OR1.19, 95%CI 1.16–1.21; 1-year all-cause OR 1.57, 95%CI 1.55–1.6; and 1-year MH/SRP OR 1.39, 95%CI 1.37–1.42). Age also influenced readmission patterns, although associations varied by outcome: higher odds of 1-year all-cause readmissions were seen with increasing age, whereas teenagers (ages 12-17) had higher odds of 1-year MH/SRP-specific readmissions than both younger children and adults. Lower odds of readmission were observed among non-White groups for most outcomes, except for 30-day readmissions, which were higher among Asian CYP (OR 1.07, 95%CI 1.03-1.11). Postpartum MH conditions were not included in analyses due to the small size in this group relative to the number of predictors.

Geographical variation was observed across England, with patients in the South West showing consistently small increased odds of readmission across all outcomes (30-day OR 1.05, 95%CI 1.01–1.09; 1-year all-cause OR 1.03, 95%CI 1.01–1.06; and 1-year MH/SRP OR 1.07, 95%CI 1.04–1.10). CYP residing in areas of higher area-level deprivation had higher odds of re-hospitalisation, with a gradient of increasing odds across higher deprivation quintiles for 1-year all-cause and MH/SRP-specific 1-year readmissions. Differences by deprivation were less pronounced for 30-day readmissions (Figure 3, Appendix S2 table ST4).

**Figure 3:**
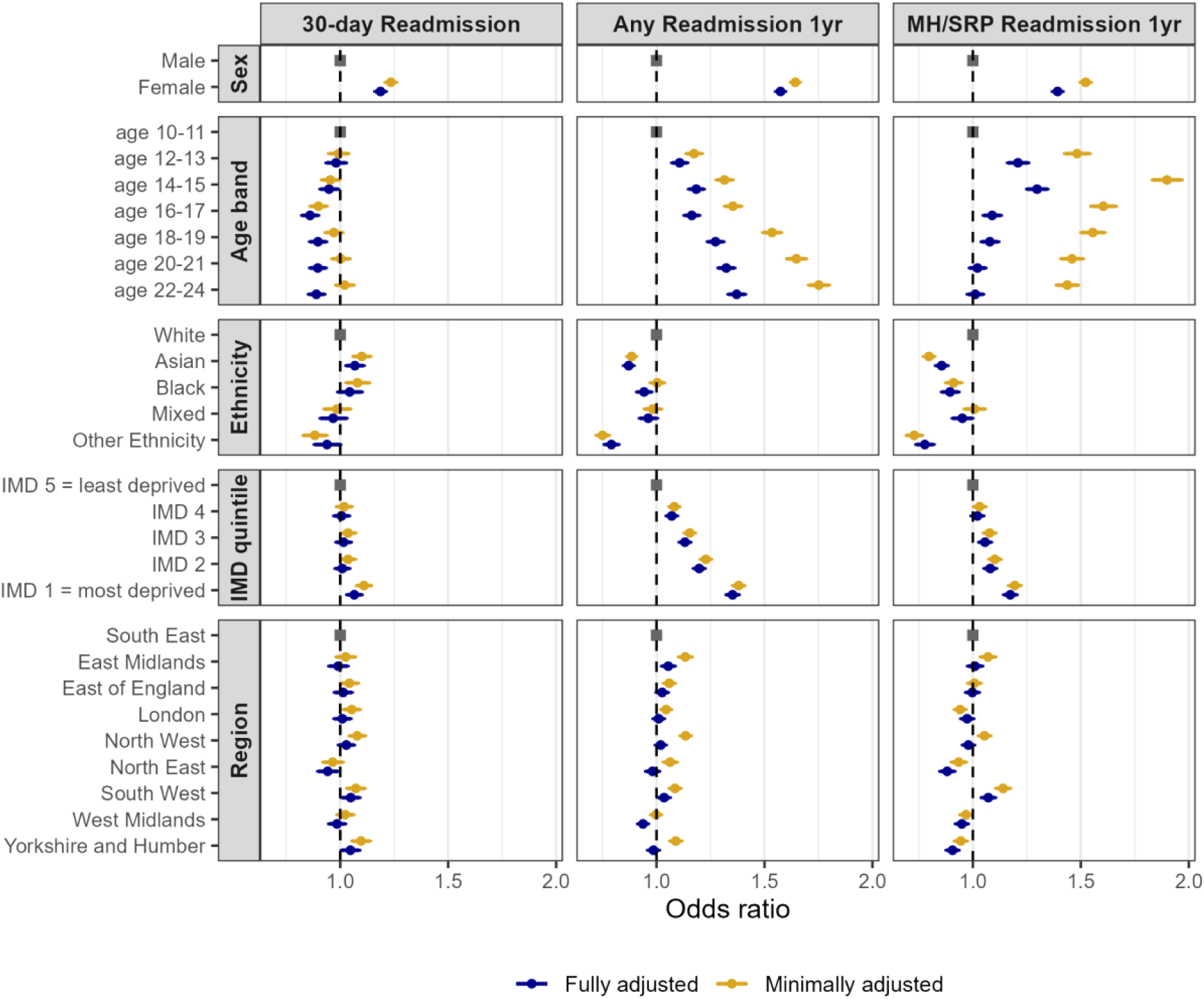
Forest plots of clinical factors associated with readmission within 30-days, 1-year and MH/SRP within 1-year. Odds ratios with 95% confidence intervals presented for fully adjusted models and minimally adjusted models (covarying for fiscal year only). Regression reference categories represented by the grey square.

Mixed results were seen for emergency vs. non-emergency admissions, with subtly higher odds of re-hospitalisation within 30 days and within 1 year for MH/SRP only (OR 1.1, 95%CI 1.07-1.13, and OR 1.1, 95%CI 1.07-1.12, respectively), but lower odds of 1-year all-cause rehospitalisation (OR 0.97, 95%CI 0.95-0.98). Self discharge was consistently associated with higher odds of rehospitalisation (30 day OR 1.08, 95%CI 1.02-1.14; 1 year all-cause OR 1.05, 95%CI 1.01-1.09; 1 year MH/SRP OR=1.04, 95%CI 1.00-1.09).

Chronic physical health conditions were associated with consistently increased odds of re-hospitalisation (30-day OR 2.67, 95%CI 2.62-2.73; 1 year all-cause OR 3.76, 95%CI 3.71-3.81; 1 year MH/SRP OR 2.32, 95%CI 2.28-2.35). Diagnostic group at index was another important predictor, with substantial variation in odds of readmission across categories relative to the reference group.

Personality disorders and eating disorders were consistently associated with higher odds of readmission, whereas substance use disorders and postpartum mental health conditions were associated with lower odds. The remaining groups exhibited more variable, outcome-specific patterns (Figure 4).

**Figure 4:**
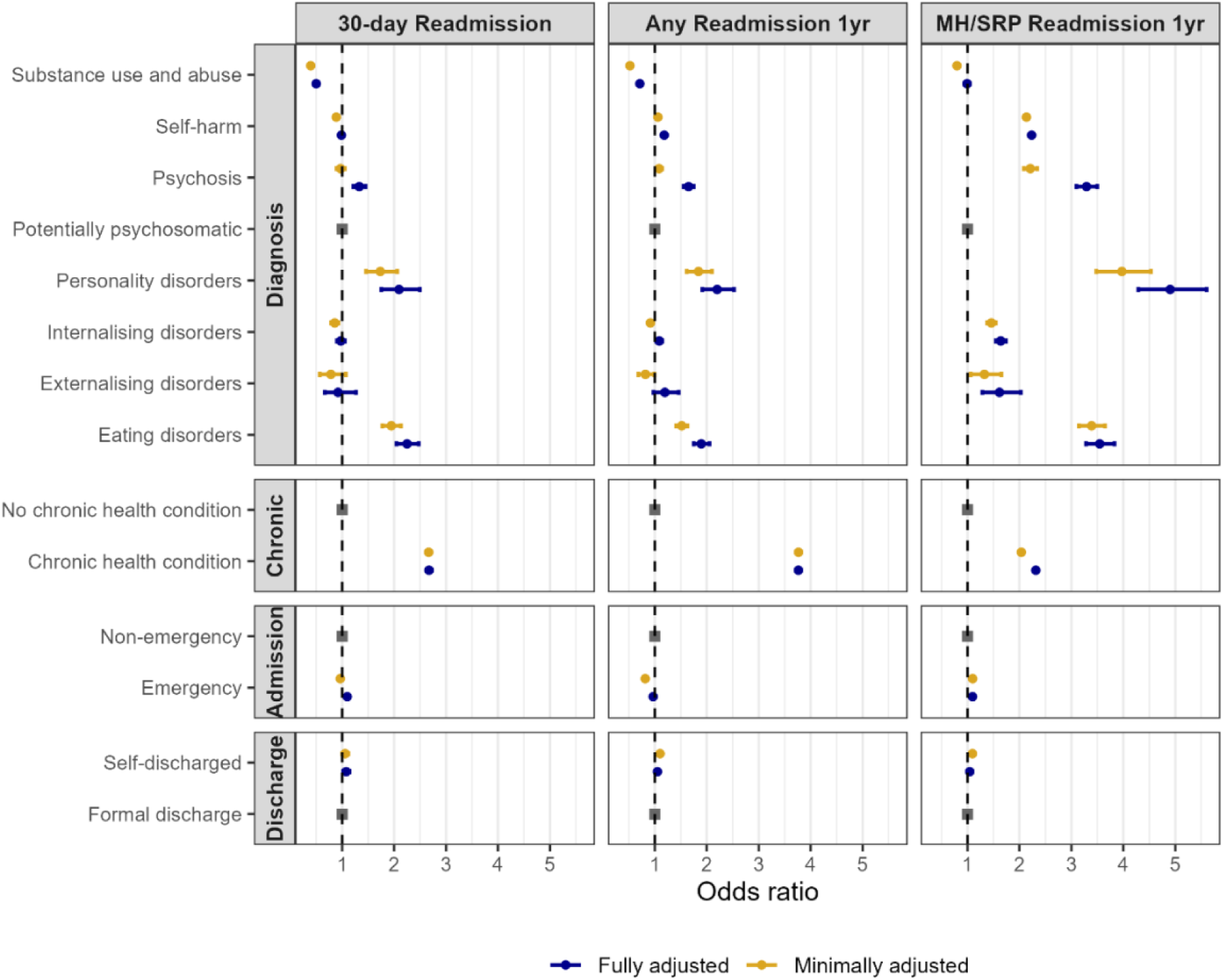
Forest plots of clinical factors associated with readmission within 30-days, 1-year and MH/SRP within 1-year. Odds ratios with 95% confidence intervals presented for fully adjusted models and minimally adjusted models (covarying for fiscal year only). Regression reference categories represented by the grey square.

### 3.5 Frequency of MH/SRP re-hospitalisations

Re-hospitalisation frequency for MH/SRP-related presentations was slightly increased in female CYP (Incidence Rate Ratio (IRR) 1.05, 95% CI 1.04–1.06), and those in mid-teenage years (age 14-15, IRR 1.03, 95%CI 1.01–1.05). Readmission frequency was lower in patients with non-white ethnicity (Asian IRR 0.97, 95%CI 0.95-0.99; Black IRR 0.95, 95%CI 0.92-0.97; Mixed IRR 0.95, 95%CI 0.920.98; Other IRR 0.93, 95%CI 0.90-0.96). IMD was not associated with frequency of re-hospitalisation.

Regionally, the highest re-hospitalisation frequency was observed in the South West (IRR 1.04, 95%CI 1.02–1.06). Diagnostic and hospital visit characteristics were associated with frequency of readmission, with the highest re-admission rates in those with chronic physical health conditions (IRR 1.2, 95% CI 1.19–1.21), and for individuals admitted for personality disorders (IRR 1.7, 95% CI 1.59–1.83), self-harm (IRR=1.3, 95%CI 1.29–1.31), and eating disorders (IRR 1.21, 95%CI 1.16–1.26). Postpartum MH conditions were not included in analyses due to the small sample size in this group compared to the number of predictors. Emergency admission (IRR=1.04, 95% CI 1.03–1.05) at index was associated with modestly higher re-hospitalisation frequency.

## 4. Discussion

This national retrospective study of MH/SRP-related hospital admissions evaluated re-hospitalisation patterns following MH/SRP admissions in CYP in England between 2014-2019. We found that re-hospitalisation was experienced by 36% CYP within 1-year of discharge. The high rate of rehospitalisation is possibly indicative of a high level of unmet need, while clinical and demographic characteristics associated with re-hospitalisation suggest that certain populations may particularly benefit from better support in the community.

### 4.1 Demographic characteristics associated with re-hospitalisation

Consistent with prior research (Ward et al., 2025), females experienced higher rates of index admissions and higher odds of re-hospitalisation. 72.4% of all CYP with at least one rehospitalisation were female. The results revealed a gradient of repeated hospitalisation with increasing deprivation, with nearly one-third of re-hospitalisations occurring among CYP living in the most deprived areas. Although effect sizes are small, these translate into large absolute differences in the context of national re-hospitalisation rates. While findings do not demonstrate interactions between gender and deprivation, they suggest that patterns of re-hospitalisation may be socially patterned along multiple, potentially overlapping axes of inequality.

Increasing age was associated with an increasing number of index admissions and re-hospitalisations, with the greatest rise in index presentations seen between ages 12-13 and 14-15. Similarly, when evaluating MH/SRP re-hospitalisations, CYP age 14-15 experienced the highest odds and frequency of re-hospitalisation in the year following their index admission. While both index admissions and readmissions were found to rise with increasing age into young adulthood, adolescence appeared to be a period of particular vulnerability, specifically for MH/SRP related re-hospitalisation.

CYP in the South West consistently experienced higher odds and frequency of readmission compared with other regions across England. These findings are in line with emergent data from the ECHILD dataset which revealed that among CYP age 11-21 in England in 2012-22, the South West had the highest levels of mental health-related hospital admissions, and lower relative use for other mental health services (Lewis, Dale, Lewer, Downs, and Blackburn, 2025). Regional differences in re-hospitalisation may reflect regional differences in the capacity, delivery and availability of services, as well as regional differences in population characteristics.

### 4.2 Clinical characteristics and diagnosis

CYP with pre-existing or co-occurring chronic physical medical health conditions were more likely to experience re-hospitalisation following an MH/SRP index admission, consistent with previous work in CYP aged 5-18 (Ward et al. 2025). Hospital visit characteristics showed modest but consistent associations with re-hospitalisation, with higher odds and frequency of re-hospitalisation seen following emergency admissions, and higher odds of rehospitalisation following self-discharge.

Diagnosis on index visit was also associated with re-hospitalisation characteristics. CYP presenting with personality disorders and eating disorders showed higher likelihood of readmission, as well as more rapid and frequent re-hospitalisation, indicating particular vulnerability in these groups. The higher risks of readmission for these diagnostic groups possibly reflects the nature of the underlying conditions. For CYP diagnosed with personality disorder, emotional crises can often escalate quickly, leading to repeat emergency hospital presentations. Psychological treatment is the cornerstone of treatment for this group, however, accessing such treatment is often difficult and is associated with long waiting lists, further increasing the likelihood of repeat hospital presentation. Eating disorders typically have a fluctuating course and medical stabilisation does not always translate into sustained recovery. For this CYP group, hospital admission may not effectively tackle psychosocial perpetuating factors, and this increases the likelihood of relapse and thus risk of repeat admission. By contrast, we found that substance use and abuse conditions were associated with the lowest odds of readmission. Although the reason for this is not clear, the pattern of results is consistent with prior meta analytic results in children under 18 years (Edgcomb et al., 2020).

### 4.3 Diagnosis at index and re-hospitalisation

For 1-year all-cause readmissions, certain diagnostic groups showed high consistency across index and successive admissions, namely for self-harm, eating disorders and psychosis. These groups have also been associated with higher rates of readmission, both within the current study and in the broader literature (Edgcomb et al., 2020).

For potentially psychosomatic presentations, nearly two thirds of subsequent re-hospitalisations were unrelated to MH/SRP (65.1%). This category was developed to capture physical manifestations of stress and distress (Geist et al., 2008), and associated admissions have been shown to rise in adolescents where schools are open and during term-time (Blackburn et al., 2021; Blackburn, Owen, Downs & Gilbert, 2022). However, the high overlap with chronic physical conditions in this group (39%), together with the lower level of subsequent mental health hospitalisation warrants further investigation.

### 4.4 Strengths and limitations

The current study uses a large, national-level administrative dataset capturing all NHS hospital admissions in England between 2014-2019. The large population sample evaluated supports age-, sex-, deprivation- and region-stratified analyses, strengthening clinical and policy-related relevance. The ability to link index admissions to readmission patterns facilitated a detailed assessment of hospital visit trajectories, allowing the capture of patterns of repeated hospital use, and the examination of timing, frequency and diagnostic continuity across successive admissions. Fine grained age-stratification allows for identification of developmental transition periods.

Evaluated data were limited to 2014-2019, precluding investigation of changes occurring during and after the pandemic. However, the data reflect a time of mandatory recording of mental health admissions data to HES APC in all hospitals, providing the data at its most compete (Villasenor et al. 2023).

The current study indicates higher odds of re-hospitalisation in those belonging to the White ethnic group compared to Black, Asian and Mixed ethnic groups, consistent with previous research (Ward et al., 2025), and prior research on health inequalities in mental healthcare (Bansal et al., 2022). However, the high rate of missing ethnicity information (around 10%), indicates that results should be interpreted with caution.

Other limitations include those inherent to HES data in particular and administrative data as a whole. Variations in diagnostic coding may have led us to underestimate the rates of rehospitalisation for specific mental health conditions (D’Sousa et al., 2024), and overestimate variability in diagnostic groupings over time. Since analyses were restricted to HES APC, the data do not provide an overview of the broader picture of mental health admissions, including access to other health services, including emergency department and outpatient services, in the period following index admissions, which may mediate re-hospitalisation statistics. Similarly, deaths recorded outside of hospital admissions were not known, leading to immortal time bias in patients who died following discharge.

## 5. Conclusion

This national retrospective study of MH/SRP-related hospital admissions among children, adolescents and young adults in England, re-hospitalisation within one year affected over one third of patients indicating substantial unmet need following discharge. Re-hospitalisation risk varied by demographic, clinical and geographic factors, with higher odds observed among females, those living in more deprived areas, adolescents and patients residing in the South West. Chronic physical health conditions, eating disorders and personality disorders were associated with more frequent and recurrent hospital use. Early-mid adolescence presents as a period of heightened vulnerability to MH/SRP-related re-hospitalisation. The findings also underscore the need for appropriately resourced, targeted, integrated and responsive services and care pathways, across all regions of the country. Bolstering such care pathways could substantially reduce the risk of repeated hospitalisation for key diagnostic groups and for those parts of the country currently less well served by robust community services.

## 6. Key points and relevance

### What’s known

Re-hospitalisation in children and young people (CYP) contributes to service pressures, may indicate gaps in care, and can disrupt education and social development.

### What’s new

Rehospitalisation following admission for mental ill-health or stress related presentations (MH/SRP) is common in England. Rehospitalisation was more likely among CYP who are female, living in deprived areas, those with cooccurring chronic physical health conditions, and particularly among CYP with eating disorders and personality disorders. Regional variation across England was also observed.

### What’s relevant

The current results have implications for clinical provision, policy, and service development. They highlight high-risk demographic and diagnostic groups, as well as regions that may benefit most from strengthened community and hospital-linked care pathways.

## Data Availability

Data can be accessed accessed via NHS Digital: https://digital.nhs.uk/services/data-accessrequest-service-dars. Analysis code can be found on GitHub.

https://github.com/ARCWest-ADS/HES_HIU/tree/main

## 7. Acknowledgements

Hospital Episode Statistics, copyright ©2024, re-used with the permission of the Health and Social Care Information Centre (“NHS Digital”). All rights reserved.

This research was supported by the National Institute for Health and Care Research (NIHR) Applied Research Collaboration (ARC) West. The views expressed in this article are those of the author(s) and not necessarily those of the NHS, the NIHR, or the Department of Health and Social Care.

AJ is supported by the NIHR Biomedical Research Centre at University Hospitals Bristol and Weston NHS Foundation Trust and the University of Bristol.

Generative AI (Chat GPT 5.1 and 5.2) was used for general editing and proofing purposes, and as a tool for R code review for this project. No data was shared with the Generative AI, and the AI did not generate, alter or manipulate any results. The manuscript represents the authors’ own work and reflects their own analyses, interpretation and insights.8. Availability of data and materials

Data in this study were provided by patients and collected by the NHS as part of their care and support. HES data can be accessed via NHS Digital: https://digital.nhs.uk/services/data-accessrequest-service-dars. CS and IW had full access to the study data and take responsibility for the integrity of the data and accuracy of the analyses. Analysis code can be found on [GitHub](https://github.com/ARCWest-ADS/HES_HIU/tree/main/analysisCode).

## Abbreviations

MH: mental health
SRP: stress-related presentations
CYP: Children and young people.

