## Supplementary material for "Repeat Hospitalisation Following Admission for Mental Ill-health and Stress-Related Presentations in Children and Young People in England between 2014-2019: A Retrospective Cohort Study": S1

### Appendix S1: ICD-10 and surgical procedure code lists

#### 1.1. Mental Health and Stress-Related Presentations

HES APC ICD-10 codes for mental health diagnostic categories and stress-related presentations, as per Ni Chobhthaigh et al. (2025). Additional codes added for Personality disorders

| **code** | **group** | **subgroup** |
| --- | --- | --- |
| F60 | personality_disorder | personality_disorder |
| F61 | personality_disorder | personality_disorder |
| F68 | personality_disorder | personality_disorder |
| F69 | personality_disorder | personality_disorder |
| F53 | postnatal_mh | postnatal_mh |
| R10 | potentially_psych | pain |
| R51 | potentially_psych | headache |
| G43 | potentially_psych | headache |
| G44 | potentially_psych | headache |
| M54 | potentially_psych | headache |
| M62.6 | potentially_psych | other pain |
| M79.6 | potentially_psych | other pain |
| M25.5 | potentially_psych | other pain |
| R52 | potentially_psych | other pain |
| R00 | potentially_psych | circulatory/respiratory signs |
| R03 | potentially_psych | circulatory/respiratory signs |
| R05 | potentially_psych | circulatory/respiratory signs |
| R06 | potentially_psych | circulatory/respiratory signs |
| R07 | potentially_psych | circulatory/respiratory signs |
| R11 | potentially_psych | digestive symptoms |
| R12 | potentially_psych | digestive symptoms |
| R13 | potentially_psych | digestive symptoms |
| R14 | potentially_psych | digestive symptoms |
| R63 | potentially_psych | digestive symptoms |
| R19.4 | potentially_psych | digestive symptoms |
| R20 | potentially_psych | skin symptoms |
| R21 | potentially_psych | skin symptoms |
| R23.1 | potentially_psych | skin symptoms |
| R23.4 | potentially_psych | skin symptoms |
| R23.8 | potentially_psych | skin symptoms |
| R25 | potentially_psych | nervous/musculoskeletal symptoms |
| R26 | potentially_psych | nervous/musculoskeletal symptoms |
| R27 | potentially_psych | nervous/musculoskeletal symptoms |
| R29.2 | potentially_psych | nervous/musculoskeletal symptoms |
| R29.3 | potentially_psych | nervous/musculoskeletal symptoms |
| R29.4 | potentially_psych | nervous/musculoskeletal symptoms |
| R29.8 | potentially_psych | nervous/musculoskeletal symptoms |
| R41 | potentially_psych | cognitive symptoms |
| R42 | potentially_psych | cognitive symptoms |
| F05 | potentially_psych | cognitive symptoms |
| R40.0 | potentially_psych | malaise/fatigue/syncope |
| R40.1 | potentially_psych | malaise/fatigue/syncope |
| R53 | potentially_psych | malaise/fatigue/syncope |
| R55 | potentially_psych | malaise/fatigue/syncope |
| N39.3 | potentially_psych | other/general symptoms |
| R44 | potentially_psych | other/general symptoms |
| R45 | potentially_psych | other/general symptoms |
| R46 | potentially_psych | other/general symptoms |
| R47 | potentially_psych | other/general symptoms |
| R49 | potentially_psych | other/general symptoms |
| Z56.3 | potentially_psych | other/general symptoms |
| Z56.4 | potentially_psych | other/general symptoms |
| Z71.1 | potentially_psych | other/general symptoms |
| Z73.3 | potentially_psych | other/general symptoms |
| F51 | potentially_psych | sleep disorders |
| G47 | potentially_psych | sleep disorders |
| F32.0 | internalising | mood disorders |
| F32.1 | internalising | mood disorders |
| F32.2 | internalising | mood disorders |
| F32.8 | internalising | mood disorders |
| F32.9 | internalising | mood disorders |
| F33.0 | internalising | mood disorders |
| F33.1 | internalising | mood disorders |
| F33.2 | internalising | mood disorders |
| F33.3 | internalising | mood disorders |
| F33.8 | internalising | mood disorders |
| F33.9 | internalising | mood disorders |
| F34.1 | internalising | mood disorders |
| F40 | internalising | anxiety/fear/oc/diss |
| F41 | internalising | anxiety/fear/oc/diss |
| F94.0 | internalising | anxiety/fear/oc/diss |
| F42 | internalising | anxiety/fear/oc/diss |
| F44 | internalising | anxiety/fear/oc/diss |
| F45 | internalising | anxiety/fear/oc/diss |
| F48 | internalising | anxiety/fear/oc/diss |
| F54 | internalising | anxiety/fear/oc/diss |
| F43 | internalising | stress/trauma |
| F62 | internalising | stress/trauma |
| F94.1 | internalising | stress/trauma |
| F94.2 | internalising | stress/trauma |
| F50 | eating_disorders | eating disorders |
| F93 | internalising | mood disorders |
| F98 | internalising | mood disorders |
| F63 | externalising | impulse_control |
| F90 | externalising | impulse_control |
| F91 | externalising | impulse_control |
| F92 | externalising | impulse_control |
| F10 | substance_use | substance_use |
| F11 | substance_use | substance_use |
| F12 | substance_use | substance_use |
| F13 | substance_use | substance_use |
| F14 | substance_use | substance_use |
| F15 | substance_use | substance_use |
| F16 | substance_use | substance_use |
| F17 | substance_use | substance_use |
| F18 | substance_use | substance_use |
| F19 | substance_use | substance_use |
| F55 | substance_use | substance_use |
| R78.0 | substance_use | substance_use |
| R78.1 | substance_use | substance_use |
| R78.2 | substance_use | substance_use |
| R78.3 | substance_use | substance_use |
| R78.4 | substance_use | substance_use |
| R78.5 | substance_use | substance_use |
| T51 | substance_use | substance_use |
| F30 | psychosis | mood_mania_psych |
| F31 | psychosis | mood_mania_psych |
| F32.3 | psychosis | mood_mania_psych |
| F20 | psychosis | schiz_or_other |
| F21 | psychosis | schiz_or_other |
| F22 | psychosis | schiz_or_other |
| F23 | psychosis | schiz_or_other |
| F24 | psychosis | schiz_or_other |
| F25 | psychosis | schiz_or_other |
| F28 | psychosis | schiz_or_other |
| F29 | psychosis | schiz_or_other |
| Y10 | selfharm | selfharm |
| Y11 | selfharm | selfharm |
| Y12 | selfharm | selfharm |
| Y13 | selfharm | selfharm |
| Y14 | selfharm | selfharm |
| Y15 | selfharm | selfharm |
| Y16 | selfharm | selfharm |
| Y17 | selfharm | selfharm |
| Y18 | selfharm | selfharm |
| Y19 | selfharm | selfharm |
| X60 | selfharm | selfharm |
| X61 | selfharm | selfharm |
| X62 | selfharm | selfharm |
| X63 | selfharm | selfharm |
| X64 | selfharm | selfharm |
| X65 | selfharm | selfharm |
| X66 | selfharm | selfharm |
| X67 | selfharm | selfharm |
| X68 | selfharm | selfharm |
| X69 | selfharm | selfharm |
| X70 | selfharm | selfharm |
| X71 | selfharm | selfharm |
| X72 | selfharm | selfharm |
| X73 | selfharm | selfharm |
| X74 | selfharm | selfharm |
| X75 | selfharm | selfharm |
| X76 | selfharm | selfharm |
| X77 | selfharm | selfharm |
| X78 | selfharm | selfharm |
| X79 | selfharm | selfharm |
| X80 | selfharm | selfharm |
| X81 | selfharm | selfharm |
| X82 | selfharm | selfharm |
| X83 | selfharm | selfharm |
| X84 | selfharm | selfharm |
| Z64.2 | selfharm | selfharm |
| Z91.5 | selfharm | selfharm |

#### 1.2. Medical exclusions for potentially psychosomatic presentations

HES APC ICD-10 codes for medical exclusions precluding diagnosis of potentially psychosomatic presentations, as per Ni Chobhthaigh et al. (2025).

| **code** | **group** | **subgroup** |
| --- | --- | --- |
| A00 | medexcl | medexcl |
| A01 | medexcl | medexcl |
| A02 | medexcl | medexcl |
| A03 | medexcl | medexcl |
| A04 | medexcl | medexcl |
| A05 | medexcl | medexcl |
| A06 | medexcl | medexcl |
| A07 | medexcl | medexcl |
| A08 | medexcl | medexcl |
| A09 | medexcl | medexcl |
| K52.0 | medexcl | medexcl |
| K52.1 | medexcl | medexcl |
| K52.9 | medexcl | medexcl |
| J09 | medexcl | medexcl |
| J10 | medexcl | medexcl |
| J13 | medexcl | medexcl |
| J14 | medexcl | medexcl |
| J15 | medexcl | medexcl |
| J16 | medexcl | medexcl |
| J17 | medexcl | medexcl |
| J18 | medexcl | medexcl |
| N81 | medexcl | medexcl |
| N83 | medexcl | medexcl |
| N85 | medexcl | medexcl |
| N86 | medexcl | medexcl |
| N87 | medexcl | medexcl |
| N88 | medexcl | medexcl |
| N89 | medexcl | medexcl |
| N90 | medexcl | medexcl |
| E28.2 | medexcl | medexcl |
| N31 | medexcl | medexcl |
| N39.0 | medexcl | medexcl |
| K55 | medexcl | medexcl |
| K56 | medexcl | medexcl |
| G40 | medexcl | medexcl |
| G41 | medexcl | medexcl |
| G45 | medexcl | medexcl |
| G46 | medexcl | medexcl |
| I60 | medexcl | medexcl |
| I61 | medexcl | medexcl |
| I62 | medexcl | medexcl |
| I63 | medexcl | medexcl |
| I64 | medexcl | medexcl |
| I65 | medexcl | medexcl |
| I66 | medexcl | medexcl |
| I67 | medexcl | medexcl |
| I68 | medexcl | medexcl |
| I69 | medexcl | medexcl |
| C15 | medexcl | medexcl |
| C16 | medexcl | medexcl |
| C17 | medexcl | medexcl |
| C18 | medexcl | medexcl |
| C19 | medexcl | medexcl |
| C20 | medexcl | medexcl |
| C21 | medexcl | medexcl |
| C22 | medexcl | medexcl |
| C23 | medexcl | medexcl |
| C24 | medexcl | medexcl |
| C25 | medexcl | medexcl |
| C26 | medexcl | medexcl |
| C30 | medexcl | medexcl |
| C31 | medexcl | medexcl |
| C32 | medexcl | medexcl |
| C33 | medexcl | medexcl |
| C34 | medexcl | medexcl |
| C37 | medexcl | medexcl |
| C38 | medexcl | medexcl |
| C39 | medexcl | medexcl |
| C40 | medexcl | medexcl |
| C41 | medexcl | medexcl |
| C45 | medexcl | medexcl |
| C46 | medexcl | medexcl |
| C47 | medexcl | medexcl |
| C48 | medexcl | medexcl |
| C49 | medexcl | medexcl |
| C51 | medexcl | medexcl |
| C52 | medexcl | medexcl |
| C53 | medexcl | medexcl |
| C54 | medexcl | medexcl |
| C55 | medexcl | medexcl |
| C56 | medexcl | medexcl |
| C57 | medexcl | medexcl |
| C58 | medexcl | medexcl |
| C64 | medexcl | medexcl |
| C65 | medexcl | medexcl |
| C66 | medexcl | medexcl |
| C67 | medexcl | medexcl |
| C68 | medexcl | medexcl |
| C69 | medexcl | medexcl |
| C70 | medexcl | medexcl |
| C71 | medexcl | medexcl |
| C72 | medexcl | medexcl |
| C73 | medexcl | medexcl |
| C74 | medexcl | medexcl |
| C75 | medexcl | medexcl |
| K35 | medexcl | medexcl |
| K36 | medexcl | medexcl |
| K37 | medexcl | medexcl |
| K38 | medexcl | medexcl |

#### 1.3. Medical exclusionary operation codes for potentially psychosomatic presentations

HES APC exclusionary operation codes precluding diagnosis of potentially psychosomatic presentations, as per Ni Chobhthaigh et al. (2025).

| **code** | **description** | **procedure_position** |
| --- | --- | --- |
| Y75.2 | laparoscopic approach to abdominal cavity nec | any |
| H01.3 | emergency excision of normal appendix | any |
| H02.9 | unspecified other excision of appendix | any |
| H01.2 | emergency excision of abnormal appendix nec | any |
| H01.9 | unspecified emergency excision of appendix | any |
| H02.1 | interval appendicectomy | any |
| H02.3 | prophylactic appendicectomy nec | any |
| H02.4 | incidental appendicectomy | any |
| H02.8 | other specified other excision of appendix | any |
| H01.1 | emergency excision of abnormal appendix and drainage hfq | any |

#### 1.4. Pregnancy-related exclusions for potentially psychosomatic presentations

HES APC ICD-10 codes for pregnancy related exclusions, as per Harron et al. (2016).

| **code** | **group** | **subgroup** |
| --- | --- | --- |
| Z37. | medexcl | medexcl |
| Z38. | medexcl | medexcl |
| O00 | medexcl | medexcl |
| O01 | medexcl | medexcl |
| O02 | medexcl | medexcl |
| O03 | medexcl | medexcl |
| O04 | medexcl | medexcl |
| O05 | medexcl | medexcl |
| O06 | medexcl | medexcl |
| O07 | medexcl | medexcl |
| O08 | medexcl | medexcl |
| P964 | medexcl | medexcl |

#### 1.5. Pregnancy-related exclusionary operation codes for potentially psychosomatic presentations

HES APC operation codes for pregnancy related exclusions, as per Harron et al. (2016).

| **code** | **description** | **procedure_position** |
| --- | --- | --- |
| R14 | Surgical induction of labour | any |
| R15 | Other induction of labour | any |
| R17 | Elective caesarean delivery | any |
| R18 | Other caesarean delivery | any |
| R19 | Breech extraction delivery | any |
| R20 | Other breech delivery | any |
| R21 | Forceps cephalic delivery | any |
| R22 | Vacuum delivery | any |
| R23 | Cephalic vaginal delivery with abnormal presentation of head | any |
| R24 | Normal delivery | any |
| R25 | Other methods of delivery | any |
| R27 | Other operations to facilitate delivery | any |

#### 1.6. Chronic medical conditions

HES APC ICD-10 codes chronic physical health conditions, adapted from Hardelid et al. (2014).

| **code** | **subgroup** | **group** |
| --- | --- | --- |
| C00 | neoplasms | chronic |
| C01 | neoplasms | chronic |
| C02 | neoplasms | chronic |
| C03 | neoplasms | chronic |
| C04 | neoplasms | chronic |
| C05 | neoplasms | chronic |
| C06 | neoplasms | chronic |
| C07 | neoplasms | chronic |
| C08 | neoplasms | chronic |
| C09 | neoplasms | chronic |
| C10 | neoplasms | chronic |
| C11 | neoplasms | chronic |
| C12 | neoplasms | chronic |
| C13 | neoplasms | chronic |
| C14 | neoplasms | chronic |
| C15 | neoplasms | chronic |
| C16 | neoplasms | chronic |
| C17 | neoplasms | chronic |
| C18 | neoplasms | chronic |
| C19 | neoplasms | chronic |
| C20 | neoplasms | chronic |
| C21 | neoplasms | chronic |
| C22 | neoplasms | chronic |
| C23 | neoplasms | chronic |
| C24 | neoplasms | chronic |
| C25 | neoplasms | chronic |
| C26 | neoplasms | chronic |
| C30 | neoplasms | chronic |
| C31 | neoplasms | chronic |
| C32 | neoplasms | chronic |
| C33 | neoplasms | chronic |
| C34 | neoplasms | chronic |
| C37 | neoplasms | chronic |
| C38 | neoplasms | chronic |
| C39 | neoplasms | chronic |
| C40 | neoplasms | chronic |
| C41 | neoplasms | chronic |
| C43 | neoplasms | chronic |
| C44 | neoplasms | chronic |
| C45 | neoplasms | chronic |
| C46 | neoplasms | chronic |
| C47 | neoplasms | chronic |
| C48 | neoplasms | chronic |
| C49 | neoplasms | chronic |
| C50 | neoplasms | chronic |
| C51 | neoplasms | chronic |
| C52 | neoplasms | chronic |
| C53 | neoplasms | chronic |
| C54 | neoplasms | chronic |
| C55 | neoplasms | chronic |
| C56 | neoplasms | chronic |
| C57 | neoplasms | chronic |
| C58 | neoplasms | chronic |
| C60 | neoplasms | chronic |
| C61 | neoplasms | chronic |
| C62 | neoplasms | chronic |
| C63 | neoplasms | chronic |
| C64 | neoplasms | chronic |
| C65 | neoplasms | chronic |
| C66 | neoplasms | chronic |
| C67 | neoplasms | chronic |
| C68 | neoplasms | chronic |
| C69 | neoplasms | chronic |
| C70 | neoplasms | chronic |
| C71 | neoplasms | chronic |
| C72 | neoplasms | chronic |
| C73 | neoplasms | chronic |
| C74 | neoplasms | chronic |
| C75 | neoplasms | chronic |
| C76 | neoplasms | chronic |
| C77 | neoplasms | chronic |
| C78 | neoplasms | chronic |
| C79 | neoplasms | chronic |
| C80 | neoplasms | chronic |
| C81 | neoplasms | chronic |
| C82 | neoplasms | chronic |
| C83 | neoplasms | chronic |
| C84 | neoplasms | chronic |
| C85 | neoplasms | chronic |
| C86 | neoplasms | chronic |
| C88 | neoplasms | chronic |
| C90 | neoplasms | chronic |
| C91 | neoplasms | chronic |
| C92 | neoplasms | chronic |
| C93 | neoplasms | chronic |
| C94 | neoplasms | chronic |
| C95 | neoplasms | chronic |
| C96 | neoplasms | chronic |
| C97 | neoplasms | chronic |
| D00 | neoplasms | chronic |
| D01 | neoplasms | chronic |
| D02 | neoplasms | chronic |
| D05 | neoplasms | chronic |
| D06 | neoplasms | chronic |
| D07 | neoplasms | chronic |
| D09 | neoplasms | chronic |
| D12 | neoplasms | chronic |
| D13 | neoplasms | chronic |
| D14.1 | neoplasms | chronic |
| D14.2 | neoplasms | chronic |
| D14.3 | neoplasms | chronic |
| D14.4 | neoplasms | chronic |
| D15 | neoplasms | chronic |
| D20 | neoplasms | chronic |
| D32 | neoplasms | chronic |
| D33 | neoplasms | chronic |
| D34 | neoplasms | chronic |
| D35 | neoplasms | chronic |
| D37 | neoplasms | chronic |
| D38 | neoplasms | chronic |
| D39 | neoplasms | chronic |
| D40 | neoplasms | chronic |
| D41 | neoplasms | chronic |
| D42 | neoplasms | chronic |
| D43 | neoplasms | chronic |
| D44 | neoplasms | chronic |
| D45 | neoplasms | chronic |
| D46 | neoplasms | chronic |
| D47 | neoplasms | chronic |
| D48 | neoplasms | chronic |
| D63.0 | neoplasms | chronic |
| E34.0 | neoplasms | chronic |
| E88.3 | neoplasms | chronic |
| G13.0 | neoplasms | chronic |
| G13.1 | neoplasms | chronic |
| G53.3 | neoplasms | chronic |
| G55.0 | neoplasms | chronic |
| G63.1 | neoplasms | chronic |
| G73.1 | neoplasms | chronic |
| G73.2 | neoplasms | chronic |
| G94.1 | neoplasms | chronic |
| M36.0 | neoplasms | chronic |
| M36.1 | neoplasms | chronic |
| M49.5 | neoplasms | chronic |
| M82.0 | neoplasms | chronic |
| M90.6 | neoplasms | chronic |
| M90.7 | neoplasms | chronic |
| N08.1 | neoplasms | chronic |
| N16.1 | neoplasms | chronic |
| Y43.1 | neoplasms | chronic |
| Y43.2 | neoplasms | chronic |
| Y43.3 | neoplasms | chronic |
| Y84.2 | neoplasms | chronic |
| Z08 | neoplasms | chronic |
| Z51.0 | neoplasms | chronic |
| Z51.1 | neoplasms | chronic |
| Z51.2 | neoplasms | chronic |
| Z54.1 | neoplasms | chronic |
| Z54.2 | neoplasms | chronic |
| Z85 | neoplasms | chronic |
| Z86.0 | neoplasms | chronic |
| Z92.3 | neoplasms | chronic |
| D80 | immunologicla disorders | chronic |
| D81 | immunologicla disorders | chronic |
| D82 | immunologicla disorders | chronic |
| D83 | immunologicla disorders | chronic |
| D84 | immunologicla disorders | chronic |
| G53.2 | immunologicla disorders | chronic |
| Q98.0 | immunologicla disorders | chronic |
| D50 | anaemia and other blood disorders | chronic |
| D56.0 | anaemia and other blood disorders | chronic |
| D56.1 | anaemia and other blood disorders | chronic |
| D56.2 | anaemia and other blood disorders | chronic |
| D56.4 | anaemia and other blood disorders | chronic |
| D56.8 | anaemia and other blood disorders | chronic |
| D56.9 | anaemia and other blood disorders | chronic |
| D57.0 | anaemia and other blood disorders | chronic |
| D57.1 | anaemia and other blood disorders | chronic |
| D57.2 | anaemia and other blood disorders | chronic |
| D57.8 | anaemia and other blood disorders | chronic |
| D58 | anaemia and other blood disorders | chronic |
| D61.0 | anaemia and other blood disorders | chronic |
| D61.9 | anaemia and other blood disorders | chronic |
| D64 | anaemia and other blood disorders | chronic |
| D66 | anaemia and other blood disorders | chronic |
| D67 | anaemia and other blood disorders | chronic |
| D68.0 | anaemia and other blood disorders | chronic |
| D68.1 | anaemia and other blood disorders | chronic |
| D68.2 | anaemia and other blood disorders | chronic |
| D68.4 | anaemia and other blood disorders | chronic |
| D68.5 | anaemia and other blood disorders | chronic |
| D68.6 | anaemia and other blood disorders | chronic |
| D68.8 | anaemia and other blood disorders | chronic |
| D68.9 | anaemia and other blood disorders | chronic |
| D69 | anaemia and other blood disorders | chronic |
| D70 | anaemia and other blood disorders | chronic |
| D71 | anaemia and other blood disorders | chronic |
| D72 | anaemia and other blood disorders | chronic |
| D73 | anaemia and other blood disorders | chronic |
| D74 | anaemia and other blood disorders | chronic |
| D75 | anaemia and other blood disorders | chronic |
| D76 | anaemia and other blood disorders | chronic |
| M36.2 | anaemia and other blood disorders | chronic |
| M36.3 | anaemia and other blood disorders | chronic |
| M36.4 | anaemia and other blood disorders | chronic |
| M90.4 | anaemia and other blood disorders | chronic |
| N08.2 | anaemia and other blood disorders | chronic |
| Z86.2 | anaemia and other blood disorders | chronic |
| B20 | hiv | chronic |
| B21 | hiv | chronic |
| B22 | hiv | chronic |
| B23 | hiv | chronic |
| B24 | hiv | chronic |
| F02.4 | hiv | chronic |
| R75 | hiv | chronic |
| Z21 | hiv | chronic |
| A15 | tuberculosis | chronic |
| A16 | tuberculosis | chronic |
| A17 | tuberculosis | chronic |
| A18 | tuberculosis | chronic |
| A19 | tuberculosis | chronic |
| E35.0 | tuberculosis | chronic |
| K23.0 | tuberculosis | chronic |
| K67.3 | tuberculosis | chronic |
| K93.0 | tuberculosis | chronic |
| M01.1 | tuberculosis | chronic |
| M49.0 | tuberculosis | chronic |
| P37.0 | tuberculosis | chronic |
| A50 | other [chronic infections] | chronic |
| A81 | other [chronic infections] | chronic |
| B18 | other [chronic infections] | chronic |
| B37.1 | other [chronic infections] | chronic |
| B37.5 | other [chronic infections] | chronic |
| B37.6 | other [chronic infections] | chronic |
| B37.7 | other [chronic infections] | chronic |
| B38.1 | other [chronic infections] | chronic |
| B39.1 | other [chronic infections] | chronic |
| B40.1 | other [chronic infections] | chronic |
| B44.0 | other [chronic infections] | chronic |
| B44.7 | other [chronic infections] | chronic |
| B45 | other [chronic infections] | chronic |
| B46 | other [chronic infections] | chronic |
| B48.7 | other [chronic infections] | chronic |
| B50.0 | other [chronic infections] | chronic |
| B50.8 | other [chronic infections] | chronic |
| B51.0 | other [chronic infections] | chronic |
| B51.8 | other [chronic infections] | chronic |
| B52.8 | other [chronic infections] | chronic |
| B52.0 | other [chronic infections] | chronic |
| B55 | other [chronic infections] | chronic |
| B57.2 | other [chronic infections] | chronic |
| B57.3 | other [chronic infections] | chronic |
| B57.4 | other [chronic infections] | chronic |
| B57.5 | other [chronic infections] | chronic |
| B58.0 | other [chronic infections] | chronic |
| B59 | other [chronic infections] | chronic |
| B67 | other [chronic infections] | chronic |
| B69 | other [chronic infections] | chronic |
| B73 | other [chronic infections] | chronic |
| B74 | other [chronic infections] | chronic |
| B78.7 | other [chronic infections] | chronic |
| B90 | other [chronic infections] | chronic |
| B91 | other [chronic infections] | chronic |
| B92 | other [chronic infections] | chronic |
| B94 | other [chronic infections] | chronic |
| F02.1 | other [chronic infections] | chronic |
| K23.1 | other [chronic infections] | chronic |
| K93.1 | other [chronic infections] | chronic |
| M00 | other [chronic infections] | chronic |
| N33.0 | other [chronic infections] | chronic |
| P35.0 | other [chronic infections] | chronic |
| P35.1 | other [chronic infections] | chronic |
| P35.2 | other [chronic infections] | chronic |
| P35.8 | other [chronic infections] | chronic |
| P35.9 | other [chronic infections] | chronic |
| P37.1 | other [chronic infections] | chronic |
| J41 | asthma and chronic lower respiratory disease | chronic |
| J42 | asthma and chronic lower respiratory disease | chronic |
| J43 | asthma and chronic lower respiratory disease | chronic |
| J44 | asthma and chronic lower respiratory disease | chronic |
| J45 | asthma and chronic lower respiratory disease | chronic |
| J46 | asthma and chronic lower respiratory disease | chronic |
| J47 | asthma and chronic lower respiratory disease | chronic |
| E84 | cystic fibrosis | chronic |
| P75 | cystic fibrosis | chronic |
| S17 | injuries | chronic |
| S27 | injuries | chronic |
| S28 | injuries | chronic |
| T27 | injuries | chronic |
| T91.4 | injuries | chronic |
| Q30 | congenital anomalies | chronic |
| Q31 | congenital anomalies | chronic |
| Q32 | congenital anomalies | chronic |
| Q33 | congenital anomalies | chronic |
| Q34 | congenital anomalies | chronic |
| Q35 | congenital anomalies | chronic |
| Q36 | congenital anomalies | chronic |
| Q37 | congenital anomalies | chronic |
| Q79.0 | congenital anomalies | chronic |
| J60 | other [respiratory] | chronic |
| J61 | other [respiratory] | chronic |
| J62 | other [respiratory] | chronic |
| J63 | other [respiratory] | chronic |
| J64 | other [respiratory] | chronic |
| J65 | other [respiratory] | chronic |
| J66 | other [respiratory] | chronic |
| J67 | other [respiratory] | chronic |
| J68 | other [respiratory] | chronic |
| J69 | other [respiratory] | chronic |
| J70 | other [respiratory] | chronic |
| J80 | other [respiratory] | chronic |
| J81 | other [respiratory] | chronic |
| J82 | other [respiratory] | chronic |
| J84 | other [respiratory] | chronic |
| J85 | other [respiratory] | chronic |
| J86 | other [respiratory] | chronic |
| J96.1 | other [respiratory] | chronic |
| J98 | other [respiratory] | chronic |
| P27 | other [respiratory] | chronic |
| Y55.6 | other [respiratory] | chronic |
| Z43.0 | other [respiratory] | chronic |
| Z93.0 | other [respiratory] | chronic |
| Z94.2 | other [respiratory] | chronic |
| E10 | diabetes | chronic |
| E11 | diabetes | chronic |
| E12 | diabetes | chronic |
| E13 | diabetes | chronic |
| E14 | diabetes | chronic |
| G59.0 | diabetes | chronic |
| G63.2 | diabetes | chronic |
| I79.2 | diabetes | chronic |
| M14.2 | diabetes | chronic |
| N08.3 | diabetes | chronic |
| O24 | diabetes | chronic |
| Y42.3 | diabetes | chronic |
| E00 | other endocrine | chronic |
| E03.0 | other endocrine | chronic |
| E03.1 | other endocrine | chronic |
| E07.1 | other endocrine | chronic |
| E22.0 | other endocrine | chronic |
| E23.0 | other endocrine | chronic |
| E25 | other endocrine | chronic |
| E26.8 | other endocrine | chronic |
| E29.1 | other endocrine | chronic |
| E31 | other endocrine | chronic |
| E34.1 | other endocrine | chronic |
| E34.2 | other endocrine | chronic |
| E34.5 | other endocrine | chronic |
| E34.8 | other endocrine | chronic |
| G13.2 | other endocrine | chronic |
| G73.5 | other endocrine | chronic |
| Y42.1 | other endocrine | chronic |
| D55 | metabolic | chronic |
| E70 | metabolic | chronic |
| E71 | metabolic | chronic |
| E72 | metabolic | chronic |
| E74 | metabolic | chronic |
| E75 | metabolic | chronic |
| E76 | metabolic | chronic |
| E77 | metabolic | chronic |
| E78 | metabolic | chronic |
| E79.1 | metabolic | chronic |
| E79.8 | metabolic | chronic |
| E79.9 | metabolic | chronic |
| E80.0 | metabolic | chronic |
| E80.1 | metabolic | chronic |
| E80.2 | metabolic | chronic |
| E80.3 | metabolic | chronic |
| E80.5 | metabolic | chronic |
| E80.7 | metabolic | chronic |
| E83 | metabolic | chronic |
| E85 | metabolic | chronic |
| E88.0 | metabolic | chronic |
| E88.1 | metabolic | chronic |
| E88.2 | metabolic | chronic |
| E88.8 | metabolic | chronic |
| E88.9 | metabolic | chronic |
| G73.6 | metabolic | chronic |
| L99.0 | metabolic | chronic |
| M14.4 | metabolic | chronic |
| M14.3 | metabolic | chronic |
| N16.3 | metabolic | chronic |
| K20 | digestive | chronic |
| K21.0 | digestive | chronic |
| K22 | digestive | chronic |
| K23.8 | digestive | chronic |
| K25 | digestive | chronic |
| K26 | digestive | chronic |
| K27 | digestive | chronic |
| K28 | digestive | chronic |
| K29.0 | digestive | chronic |
| K29.1 | digestive | chronic |
| K29.3 | digestive | chronic |
| K29.4 | digestive | chronic |
| K29.5 | digestive | chronic |
| K29.6 | digestive | chronic |
| K29.7 | digestive | chronic |
| K29.8 | digestive | chronic |
| K29.9 | digestive | chronic |
| K31 | digestive | chronic |
| K50 | digestive | chronic |
| K51 | digestive | chronic |
| K52 | digestive | chronic |
| K55 | digestive | chronic |
| K57 | digestive | chronic |
| K59.2 | digestive | chronic |
| K63.0 | digestive | chronic |
| K63.1 | digestive | chronic |
| K63.2 | digestive | chronic |
| K63.3 | digestive | chronic |
| K66 | digestive | chronic |
| K72 | digestive | chronic |
| K73 | digestive | chronic |
| K74 | digestive | chronic |
| K75 | digestive | chronic |
| K76 | digestive | chronic |
| K80 | digestive | chronic |
| K81 | digestive | chronic |
| K82 | digestive | chronic |
| K83 | digestive | chronic |
| K85.0 | digestive | chronic |
| K85.1 | digestive | chronic |
| K85.8 | digestive | chronic |
| K85.9 | digestive | chronic |
| K86.1 | digestive | chronic |
| K86.2 | digestive | chronic |
| K86.3 | digestive | chronic |
| K86.8 | digestive | chronic |
| K86.9 | digestive | chronic |
| K87.0 | digestive | chronic |
| K90 | digestive | chronic |
| M07.4 | digestive | chronic |
| M07.5 | digestive | chronic |
| M09.1 | digestive | chronic |
| M09.2 | digestive | chronic |
| T86.4 | digestive | chronic |
| Z43.2 | digestive | chronic |
| Z43.3 | digestive | chronic |
| Z43.4 | digestive | chronic |
| Z46.5 | digestive | chronic |
| Z90.3 | digestive | chronic |
| Z90.4 | digestive | chronic |
| Z93.2 | digestive | chronic |
| Z93.3 | digestive | chronic |
| Z93.4 | digestive | chronic |
| Z93.5 | digestive | chronic |
| D63.8 | renal/gu | chronic |
| G63.8 | renal/gu | chronic |
| G99.8 | renal/gu | chronic |
| I68.8 | renal/gu | chronic |
| M90.8 | renal/gu | chronic |
| N08.4 | renal/gu | chronic |
| N00 | renal/gu | chronic |
| N01 | renal/gu | chronic |
| N02 | renal/gu | chronic |
| N03 | renal/gu | chronic |
| N04 | renal/gu | chronic |
| N05 | renal/gu | chronic |
| N07 | renal/gu | chronic |
| N11 | renal/gu | chronic |
| N12 | renal/gu | chronic |
| N13 | renal/gu | chronic |
| N14 | renal/gu | chronic |
| N15 | renal/gu | chronic |
| N16.0 | renal/gu | chronic |
| N16.2 | renal/gu | chronic |
| N16.4 | renal/gu | chronic |
| N16.5 | renal/gu | chronic |
| N16.8 | renal/gu | chronic |
| N18 | renal/gu | chronic |
| N19 | renal/gu | chronic |
| N20 | renal/gu | chronic |
| N21 | renal/gu | chronic |
| N22 | renal/gu | chronic |
| N23 | renal/gu | chronic |
| N25 | renal/gu | chronic |
| N26 | renal/gu | chronic |
| N28 | renal/gu | chronic |
| N29 | renal/gu | chronic |
| N31 | renal/gu | chronic |
| N32 | renal/gu | chronic |
| N33.8 | renal/gu | chronic |
| N35 | renal/gu | chronic |
| N36 | renal/gu | chronic |
| N39.1 | renal/gu | chronic |
| N39.4 | renal/gu | chronic |
| N40 | renal/gu | chronic |
| N41 | renal/gu | chronic |
| N42 | renal/gu | chronic |
| N70 | renal/gu | chronic |
| N71 | renal/gu | chronic |
| N72 | renal/gu | chronic |
| N73 | renal/gu | chronic |
| N74 | renal/gu | chronic |
| N80 | renal/gu | chronic |
| N81 | renal/gu | chronic |
| N82 | renal/gu | chronic |
| N85 | renal/gu | chronic |
| N86 | renal/gu | chronic |
| N87 | renal/gu | chronic |
| N88 | renal/gu | chronic |
| P96.0 | renal/gu | chronic |
| T82.4 | renal/gu | chronic |
| T83.1 | renal/gu | chronic |
| T83.2 | renal/gu | chronic |
| T83.4 | renal/gu | chronic |
| T83.5 | renal/gu | chronic |
| T83.6 | renal/gu | chronic |
| T83.8 | renal/gu | chronic |
| T83.9 | renal/gu | chronic |
| T85.5 | renal/gu | chronic |
| T86.1 | renal/gu | chronic |
| Y60.2 | renal/gu | chronic |
| Y61.2 | renal/gu | chronic |
| Y62.2 | renal/gu | chronic |
| Y84.1 | renal/gu | chronic |
| Z49 | renal/gu | chronic |
| Z93.6 | renal/gu | chronic |
| Z94.0 | renal/gu | chronic |
| Z99.2 | renal/gu | chronic |
| Q38.0 | congenital anomalies of the digestive/renal/gu system | chronic |
| Q38.3 | congenital anomalies of the digestive/renal/gu system | chronic |
| Q38.4 | congenital anomalies of the digestive/renal/gu system | chronic |
| Q38.6 | congenital anomalies of the digestive/renal/gu system | chronic |
| Q38.7 | congenital anomalies of the digestive/renal/gu system | chronic |
| Q38.8 | congenital anomalies of the digestive/renal/gu system | chronic |
| Q39 | congenital anomalies of the digestive/renal/gu system | chronic |
| Q40.2 | congenital anomalies of the digestive/renal/gu system | chronic |
| Q40.3 | congenital anomalies of the digestive/renal/gu system | chronic |
| Q40.8 | congenital anomalies of the digestive/renal/gu system | chronic |
| Q40.9 | congenital anomalies of the digestive/renal/gu system | chronic |
| Q41 | congenital anomalies of the digestive/renal/gu system | chronic |
| Q42 | congenital anomalies of the digestive/renal/gu system | chronic |
| Q43.1 | congenital anomalies of the digestive/renal/gu system | chronic |
| Q43.3 | congenital anomalies of the digestive/renal/gu system | chronic |
| Q43.4 | congenital anomalies of the digestive/renal/gu system | chronic |
| Q43.5 | congenital anomalies of the digestive/renal/gu system | chronic |
| Q43.6 | congenital anomalies of the digestive/renal/gu system | chronic |
| Q43.7 | congenital anomalies of the digestive/renal/gu system | chronic |
| Q43.9 | congenital anomalies of the digestive/renal/gu system | chronic |
| Q44 | congenital anomalies of the digestive/renal/gu system | chronic |
| Q45 | congenital anomalies of the digestive/renal/gu system | chronic |
| Q50.0 | congenital anomalies of the digestive/renal/gu system | chronic |
| Q51 | congenital anomalies of the digestive/renal/gu system | chronic |
| Q52.0 | congenital anomalies of the digestive/renal/gu system | chronic |
| Q52.1 | congenital anomalies of the digestive/renal/gu system | chronic |
| Q52.2 | congenital anomalies of the digestive/renal/gu system | chronic |
| Q52.4 | congenital anomalies of the digestive/renal/gu system | chronic |
| Q54.0 | congenital anomalies of the digestive/renal/gu system | chronic |
| Q54.1 | congenital anomalies of the digestive/renal/gu system | chronic |
| Q54.2 | congenital anomalies of the digestive/renal/gu system | chronic |
| Q54.3 | congenital anomalies of the digestive/renal/gu system | chronic |
| Q54.8 | congenital anomalies of the digestive/renal/gu system | chronic |
| Q54.9 | congenital anomalies of the digestive/renal/gu system | chronic |
| Q55.0 | congenital anomalies of the digestive/renal/gu system | chronic |
| Q55.5 | congenital anomalies of the digestive/renal/gu system | chronic |
| Q56 | congenital anomalies of the digestive/renal/gu system | chronic |
| Q60.1 | congenital anomalies of the digestive/renal/gu system | chronic |
| Q60.2 | congenital anomalies of the digestive/renal/gu system | chronic |
| Q60.4 | congenital anomalies of the digestive/renal/gu system | chronic |
| Q60.5 | congenital anomalies of the digestive/renal/gu system | chronic |
| Q60.6 | congenital anomalies of the digestive/renal/gu system | chronic |
| Q61 | congenital anomalies of the digestive/renal/gu system | chronic |
| Q62.0 | congenital anomalies of the digestive/renal/gu system | chronic |
| Q62.1 | congenital anomalies of the digestive/renal/gu system | chronic |
| Q62.2 | congenital anomalies of the digestive/renal/gu system | chronic |
| Q62.3 | congenital anomalies of the digestive/renal/gu system | chronic |
| Q62.4 | congenital anomalies of the digestive/renal/gu system | chronic |
| Q62.5 | congenital anomalies of the digestive/renal/gu system | chronic |
| Q62.6 | congenital anomalies of the digestive/renal/gu system | chronic |
| Q62.8 | congenital anomalies of the digestive/renal/gu system | chronic |
| Q63.0 | congenital anomalies of the digestive/renal/gu system | chronic |
| Q63.1 | congenital anomalies of the digestive/renal/gu system | chronic |
| Q63.2 | congenital anomalies of the digestive/renal/gu system | chronic |
| Q63.8 | congenital anomalies of the digestive/renal/gu system | chronic |
| Q63.9 | congenital anomalies of the digestive/renal/gu system | chronic |
| Q64 | congenital anomalies of the digestive/renal/gu system | chronic |
| Q79.2 | congenital anomalies of the digestive/renal/gu system | chronic |
| Q79.3 | congenital anomalies of the digestive/renal/gu system | chronic |
| Q79.4 | congenital anomalies of the digestive/renal/gu system | chronic |
| Q79.5 | congenital anomalies of the digestive/renal/gu system | chronic |
| Q87.8 | congenital anomalies of the digestive/renal/gu system | chronic |
| Q89.1 | congenital anomalies of the digestive/renal/gu system | chronic |
| Q89.2 | congenital anomalies of the digestive/renal/gu system | chronic |
| S36 | injuries | chronic |
| S37 | injuries | chronic |
| S38 | injuries | chronic |
| S39.6 | injuries | chronic |
| S39.7 | injuries | chronic |
| T06.5 | injuries | chronic |
| T28 | injuries | chronic |
| T91.5 | injuries | chronic |
| E66 | other/unspecific [metabolic etc] | chronic |
| G63.3 | other/unspecific [metabolic etc] | chronic |
| G99.0 | other/unspecific [metabolic etc] | chronic |
| M14.5 | other/unspecific [metabolic etc] | chronic |
| N92 | other/unspecific [metabolic etc] | chronic |
| Z86.3 | other/unspecific [metabolic etc] | chronic |
| Z93.8 | other/unspecific [metabolic etc] | chronic |
| G55.1 | musculoskeletal/connective tissue | chronic |
| G55.2 | musculoskeletal/connective tissue | chronic |
| G55.3 | musculoskeletal/connective tissue | chronic |
| G63.5 | musculoskeletal/connective tissue | chronic |
| G63.6 | musculoskeletal/connective tissue | chronic |
| G73.7 | musculoskeletal/connective tissue | chronic |
| J99.0 | musculoskeletal/connective tissue | chronic |
| J99.1 | musculoskeletal/connective tissue | chronic |
| L62.0 | musculoskeletal/connective tissue | chronic |
| M05 | musculoskeletal/connective tissue | chronic |
| M06 | musculoskeletal/connective tissue | chronic |
| M07.0 | musculoskeletal/connective tissue | chronic |
| M07.1 | musculoskeletal/connective tissue | chronic |
| M07.2 | musculoskeletal/connective tissue | chronic |
| M07.3 | musculoskeletal/connective tissue | chronic |
| M07.6 | musculoskeletal/connective tissue | chronic |
| M08 | musculoskeletal/connective tissue | chronic |
| M09.8 | musculoskeletal/connective tissue | chronic |
| M10 | musculoskeletal/connective tissue | chronic |
| M11 | musculoskeletal/connective tissue | chronic |
| M12 | musculoskeletal/connective tissue | chronic |
| M13 | musculoskeletal/connective tissue | chronic |
| M14.0 | musculoskeletal/connective tissue | chronic |
| M14.6 | musculoskeletal/connective tissue | chronic |
| M14.8 | musculoskeletal/connective tissue | chronic |
| M30 | musculoskeletal/connective tissue | chronic |
| M31 | musculoskeletal/connective tissue | chronic |
| M32 | musculoskeletal/connective tissue | chronic |
| M33 | musculoskeletal/connective tissue | chronic |
| M34 | musculoskeletal/connective tissue | chronic |
| M35 | musculoskeletal/connective tissue | chronic |
| M40 | musculoskeletal/connective tissue | chronic |
| M41 | musculoskeletal/connective tissue | chronic |
| M42 | musculoskeletal/connective tissue | chronic |
| M43 | musculoskeletal/connective tissue | chronic |
| M45 | musculoskeletal/connective tissue | chronic |
| M46 | musculoskeletal/connective tissue | chronic |
| M47 | musculoskeletal/connective tissue | chronic |
| M48 | musculoskeletal/connective tissue | chronic |
| M50 | musculoskeletal/connective tissue | chronic |
| M51 | musculoskeletal/connective tissue | chronic |
| M53 | musculoskeletal/connective tissue | chronic |
| M60 | musculoskeletal/connective tissue | chronic |
| M61 | musculoskeletal/connective tissue | chronic |
| M62 | musculoskeletal/connective tissue | chronic |
| M63.8 | musculoskeletal/connective tissue | chronic |
| M80.1 | musculoskeletal/connective tissue | chronic |
| M80.2 | musculoskeletal/connective tissue | chronic |
| M80.3 | musculoskeletal/connective tissue | chronic |
| M80.4 | musculoskeletal/connective tissue | chronic |
| M80.5 | musculoskeletal/connective tissue | chronic |
| M80.8 | musculoskeletal/connective tissue | chronic |
| M80.9 | musculoskeletal/connective tissue | chronic |
| M81.1 | musculoskeletal/connective tissue | chronic |
| M81.2 | musculoskeletal/connective tissue | chronic |
| M81.3 | musculoskeletal/connective tissue | chronic |
| M81.4 | musculoskeletal/connective tissue | chronic |
| M81.5 | musculoskeletal/connective tissue | chronic |
| M81.6 | musculoskeletal/connective tissue | chronic |
| M81.8 | musculoskeletal/connective tissue | chronic |
| M81.9 | musculoskeletal/connective tissue | chronic |
| M82.1 | musculoskeletal/connective tissue | chronic |
| M82.8 | musculoskeletal/connective tissue | chronic |
| M84.0 | musculoskeletal/connective tissue | chronic |
| M84.1 | musculoskeletal/connective tissue | chronic |
| M84.2 | musculoskeletal/connective tissue | chronic |
| M84.8 | musculoskeletal/connective tissue | chronic |
| M84.9 | musculoskeletal/connective tissue | chronic |
| M85 | musculoskeletal/connective tissue | chronic |
| M86.3 | musculoskeletal/connective tissue | chronic |
| M86.4 | musculoskeletal/connective tissue | chronic |
| M86.5 | musculoskeletal/connective tissue | chronic |
| M86.6 | musculoskeletal/connective tissue | chronic |
| M89 | musculoskeletal/connective tissue | chronic |
| M90.0 | musculoskeletal/connective tissue | chronic |
| M91 | musculoskeletal/connective tissue | chronic |
| M92 | musculoskeletal/connective tissue | chronic |
| M93 | musculoskeletal/connective tissue | chronic |
| M94 | musculoskeletal/connective tissue | chronic |
| N08.5 | musculoskeletal/connective tissue | chronic |
| Y45.4 | musculoskeletal/connective tissue | chronic |
| S13 | skeletal injuries/amputations | chronic |
| S22.0 | skeletal injuries/amputations | chronic |
| S22.1 | skeletal injuries/amputations | chronic |
| S22.2 | skeletal injuries/amputations | chronic |
| S22.5 | skeletal injuries/amputations | chronic |
| S23 | skeletal injuries/amputations | chronic |
| S32 | skeletal injuries/amputations | chronic |
| S33 | skeletal injuries/amputations | chronic |
| S68.3 | skeletal injuries/amputations | chronic |
| S68.4 | skeletal injuries/amputations | chronic |
| S68.8 | skeletal injuries/amputations | chronic |
| S77 | skeletal injuries/amputations | chronic |
| S78 | skeletal injuries/amputations | chronic |
| S87 | skeletal injuries/amputations | chronic |
| S88 | skeletal injuries/amputations | chronic |
| S97 | skeletal injuries/amputations | chronic |
| S98.0 | skeletal injuries/amputations | chronic |
| S98.2 | skeletal injuries/amputations | chronic |
| S98.3 | skeletal injuries/amputations | chronic |
| S98.4 | skeletal injuries/amputations | chronic |
| T02 | skeletal injuries/amputations | chronic |
| T04 | skeletal injuries/amputations | chronic |
| T05 | skeletal injuries/amputations | chronic |
| T20.3 | skeletal injuries/amputations | chronic |
| T20.7 | skeletal injuries/amputations | chronic |
| T21.3 | skeletal injuries/amputations | chronic |
| T21.7 | skeletal injuries/amputations | chronic |
| T22.3 | skeletal injuries/amputations | chronic |
| T22.7 | skeletal injuries/amputations | chronic |
| T23.2 | skeletal injuries/amputations | chronic |
| T23.3 | skeletal injuries/amputations | chronic |
| T23.6 | skeletal injuries/amputations | chronic |
| T23.7 | skeletal injuries/amputations | chronic |
| T24.3 | skeletal injuries/amputations | chronic |
| T24.7 | skeletal injuries/amputations | chronic |
| T25.2 | skeletal injuries/amputations | chronic |
| T25.3 | skeletal injuries/amputations | chronic |
| T25.6 | skeletal injuries/amputations | chronic |
| T25.7 | skeletal injuries/amputations | chronic |
| T29.3 | skeletal injuries/amputations | chronic |
| T29.7 | skeletal injuries/amputations | chronic |
| T30.3 | skeletal injuries/amputations | chronic |
| T30.7 | skeletal injuries/amputations | chronic |
| T31.2 | skeletal injuries/amputations | chronic |
| T31.3 | skeletal injuries/amputations | chronic |
| T31.4 | skeletal injuries/amputations | chronic |
| T31.5 | skeletal injuries/amputations | chronic |
| T31.6 | skeletal injuries/amputations | chronic |
| T31.7 | skeletal injuries/amputations | chronic |
| T31.8 | skeletal injuries/amputations | chronic |
| T31.9 | skeletal injuries/amputations | chronic |
| T32.2 | skeletal injuries/amputations | chronic |
| T32.3 | skeletal injuries/amputations | chronic |
| T32.4 | skeletal injuries/amputations | chronic |
| T32.5 | skeletal injuries/amputations | chronic |
| T32.6 | skeletal injuries/amputations | chronic |
| T32.7 | skeletal injuries/amputations | chronic |
| T32.8 | skeletal injuries/amputations | chronic |
| T32.9 | skeletal injuries/amputations | chronic |
| T87.3 | skeletal injuries/amputations | chronic |
| T87.4 | skeletal injuries/amputations | chronic |
| T87.5 | skeletal injuries/amputations | chronic |
| T87.6 | skeletal injuries/amputations | chronic |
| T91.2 | skeletal injuries/amputations | chronic |
| T91.8 | skeletal injuries/amputations | chronic |
| T92.6 | skeletal injuries/amputations | chronic |
| T93.1 | skeletal injuries/amputations | chronic |
| T93.4 | skeletal injuries/amputations | chronic |
| T93.6 | skeletal injuries/amputations | chronic |
| T94.0 | skeletal injuries/amputations | chronic |
| T94.1 | skeletal injuries/amputations | chronic |
| T95.0 | skeletal injuries/amputations | chronic |
| T95.1 | skeletal injuries/amputations | chronic |
| T95.4 | skeletal injuries/amputations | chronic |
| T95.8 | skeletal injuries/amputations | chronic |
| T95.9 | skeletal injuries/amputations | chronic |
| Y83.5 | skeletal injuries/amputations | chronic |
| Z89.1 | skeletal injuries/amputations | chronic |
| Z89.2 | skeletal injuries/amputations | chronic |
| Z89.5 | skeletal injuries/amputations | chronic |
| Z89.6 | skeletal injuries/amputations | chronic |
| Z89.7 | skeletal injuries/amputations | chronic |
| Z89.8 | skeletal injuries/amputations | chronic |
| Z97.1 | skeletal injuries/amputations | chronic |
| L10 | chronic skin disorders | chronic |
| L11.0 | chronic skin disorders | chronic |
| L11.8 | chronic skin disorders | chronic |
| L11.9 | chronic skin disorders | chronic |
| L12 | chronic skin disorders | chronic |
| L13 | chronic skin disorders | chronic |
| L14 | chronic skin disorders | chronic |
| L28 | chronic skin disorders | chronic |
| L40 | chronic skin disorders | chronic |
| L41 | chronic skin disorders | chronic |
| L42 | chronic skin disorders | chronic |
| L43 | chronic skin disorders | chronic |
| L44 | chronic skin disorders | chronic |
| L45 | chronic skin disorders | chronic |
| L57 | chronic skin disorders | chronic |
| L58.1 | chronic skin disorders | chronic |
| L59 | chronic skin disorders | chronic |
| L87 | chronic skin disorders | chronic |
| L88 | chronic skin disorders | chronic |
| L90 | chronic skin disorders | chronic |
| L92 | chronic skin disorders | chronic |
| L95 | chronic skin disorders | chronic |
| L93 | chronic skin disorders | chronic |
| L98.5 | chronic skin disorders | chronic |
| M09.0 | chronic skin disorders | chronic |
| Q80 | chronic skin disorders | chronic |
| Q81 | chronic skin disorders | chronic |
| Q87.0 | chronic skin disorders | chronic |
| Q87.1 | chronic skin disorders | chronic |
| Q87.2 | chronic skin disorders | chronic |
| Q87.3 | chronic skin disorders | chronic |
| Q87.4 | chronic skin disorders | chronic |
| Q87.5 | chronic skin disorders | chronic |
| Q89.4 | chronic skin disorders | chronic |
| Q18.8 | congenital anomalies | chronic |
| Q65.0 | congenital anomalies | chronic |
| Q65.1 | congenital anomalies | chronic |
| Q65.2 | congenital anomalies | chronic |
| Q65.8 | congenital anomalies | chronic |
| Q65.9 | congenital anomalies | chronic |
| Q67.5 | congenital anomalies | chronic |
| Q68.2 | congenital anomalies | chronic |
| Q68.3 | congenital anomalies | chronic |
| Q68.4 | congenital anomalies | chronic |
| Q68.5 | congenital anomalies | chronic |
| Q71 | congenital anomalies | chronic |
| Q72 | congenital anomalies | chronic |
| Q73 | congenital anomalies | chronic |
| Q74 | congenital anomalies | chronic |
| Q75.3 | congenital anomalies | chronic |
| Q75.4 | congenital anomalies | chronic |
| Q75.5 | congenital anomalies | chronic |
| Q75.8 | congenital anomalies | chronic |
| Q75.9 | congenital anomalies | chronic |
| Q76.1 | congenital anomalies | chronic |
| Q76.2 | congenital anomalies | chronic |
| Q76.3 | congenital anomalies | chronic |
| Q76.4 | congenital anomalies | chronic |
| Q77 | congenital anomalies | chronic |
| Q78 | congenital anomalies | chronic |
| Q79.6 | congenital anomalies | chronic |
| Q79.8 | congenital anomalies | chronic |
| Q82.0 | congenital anomalies | chronic |
| Q82.1 | congenital anomalies | chronic |
| Q82.2 | congenital anomalies | chronic |
| Q82.3 | congenital anomalies | chronic |
| Q82.4 | congenital anomalies | chronic |
| Q82.9 | congenital anomalies | chronic |
| Q86.2 | congenital anomalies | chronic |
| Q89.7 | congenital anomalies | chronic |
| Q89.8 | congenital anomalies | chronic |
| Q89.9 | congenital anomalies | chronic |
| F80.3 | epilepsy | chronic |
| G40.0 | epilepsy | chronic |
| G40.1 | epilepsy | chronic |
| G40.2 | epilepsy | chronic |
| G40.3 | epilepsy | chronic |
| G40.4 | epilepsy | chronic |
| G40.6 | epilepsy | chronic |
| G40.7 | epilepsy | chronic |
| G40.8 | epilepsy | chronic |
| G40.9 | epilepsy | chronic |
| G41 | epilepsy | chronic |
| R56.8 | epilepsy | chronic |
| Y46.0 | epilepsy | chronic |
| Y46.1 | epilepsy | chronic |
| Y46.2 | epilepsy | chronic |
| Y46.3 | epilepsy | chronic |
| Y46.4 | epilepsy | chronic |
| Y46.5 | epilepsy | chronic |
| Y46.6 | epilepsy | chronic |
| G80 | cerebral palsy | chronic |
| G81 | cerebral palsy | chronic |
| G82 | cerebral palsy | chronic |
| G83 | cerebral palsy | chronic |
| S05 | injuries of brain nerves eyes or ears | chronic |
| S06 | injuries of brain nerves eyes or ears | chronic |
| S07 | injuries of brain nerves eyes or ears | chronic |
| S08 | injuries of brain nerves eyes or ears | chronic |
| S12 | injuries of brain nerves eyes or ears | chronic |
| S14 | injuries of brain nerves eyes or ears | chronic |
| S24 | injuries of brain nerves eyes or ears | chronic |
| S34 | injuries of brain nerves eyes or ears | chronic |
| S44 | injuries of brain nerves eyes or ears | chronic |
| S54 | injuries of brain nerves eyes or ears | chronic |
| S64 | injuries of brain nerves eyes or ears | chronic |
| S74 | injuries of brain nerves eyes or ears | chronic |
| S84 | injuries of brain nerves eyes or ears | chronic |
| S94 | injuries of brain nerves eyes or ears | chronic |
| T06.0 | injuries of brain nerves eyes or ears | chronic |
| T06.1 | injuries of brain nerves eyes or ears | chronic |
| T06.2 | injuries of brain nerves eyes or ears | chronic |
| T26 | injuries of brain nerves eyes or ears | chronic |
| T90.4 | injuries of brain nerves eyes or ears | chronic |
| T90.5 | injuries of brain nerves eyes or ears | chronic |
| T91.1 | injuries of brain nerves eyes or ears | chronic |
| T91.3 | injuries of brain nerves eyes or ears | chronic |
| T92.4 | injuries of brain nerves eyes or ears | chronic |
| H05.1 | chronic eye conditions | chronic |
| H05.2 | chronic eye conditions | chronic |
| H05.3 | chronic eye conditions | chronic |
| H05.4 | chronic eye conditions | chronic |
| H05.5 | chronic eye conditions | chronic |
| H05.8 | chronic eye conditions | chronic |
| H05.9 | chronic eye conditions | chronic |
| H13.3 | chronic eye conditions | chronic |
| H17 | chronic eye conditions | chronic |
| H18 | chronic eye conditions | chronic |
| H19.3 | chronic eye conditions | chronic |
| H19.8 | chronic eye conditions | chronic |
| H21 | chronic eye conditions | chronic |
| H26 | chronic eye conditions | chronic |
| H27 | chronic eye conditions | chronic |
| H28.0 | chronic eye conditions | chronic |
| H28.1 | chronic eye conditions | chronic |
| H28.2 | chronic eye conditions | chronic |
| H31 | chronic eye conditions | chronic |
| H32.8 | chronic eye conditions | chronic |
| H33 | chronic eye conditions | chronic |
| H34 | chronic eye conditions | chronic |
| H35 | chronic eye conditions | chronic |
| H40 | chronic eye conditions | chronic |
| H42.0 | chronic eye conditions | chronic |
| H43 | chronic eye conditions | chronic |
| H44 | chronic eye conditions | chronic |
| H47 | chronic eye conditions | chronic |
| H54.0 | chronic eye conditions | chronic |
| H54.1 | chronic eye conditions | chronic |
| H54.2 | chronic eye conditions | chronic |
| H54.4 | chronic eye conditions | chronic |
| T85.2 | chronic eye conditions | chronic |
| T85.3 | chronic eye conditions | chronic |
| Z44.2 | chronic eye conditions | chronic |
| H60.2 | chronic ear conditions | chronic |
| H65.2 | chronic ear conditions | chronic |
| H65.3 | chronic ear conditions | chronic |
| H65.4 | chronic ear conditions | chronic |
| H66.1 | chronic ear conditions | chronic |
| H66.2 | chronic ear conditions | chronic |
| H66.3 | chronic ear conditions | chronic |
| H69.0 | chronic ear conditions | chronic |
| H70.1 | chronic ear conditions | chronic |
| H73.1 | chronic ear conditions | chronic |
| H74.0 | chronic ear conditions | chronic |
| H74.1 | chronic ear conditions | chronic |
| H74.2 | chronic ear conditions | chronic |
| H74.3 | chronic ear conditions | chronic |
| H75.0 | chronic ear conditions | chronic |
| H80 | chronic ear conditions | chronic |
| H81.0 | chronic ear conditions | chronic |
| H81.4 | chronic ear conditions | chronic |
| H83.0 | chronic ear conditions | chronic |
| H83.2 | chronic ear conditions | chronic |
| H90.0 | chronic ear conditions | chronic |
| H90.3 | chronic ear conditions | chronic |
| H90.5 | chronic ear conditions | chronic |
| H90.6 | chronic ear conditions | chronic |
| H91 | chronic ear conditions | chronic |
| Z45.3 | chronic ear conditions | chronic |
| P10 | perinatal conditions | chronic |
| P21.0 | perinatal conditions | chronic |
| P52 | perinatal conditions | chronic |
| P57 | perinatal conditions | chronic |
| P90 | perinatal conditions | chronic |
| P91.1 | perinatal conditions | chronic |
| P91.2 | perinatal conditions | chronic |
| P91.6 | perinatal conditions | chronic |
| Q00 | congenital anomalies of neurological or sensory systems | chronic |
| Q01 | congenital anomalies of neurological or sensory systems | chronic |
| Q02 | congenital anomalies of neurological or sensory systems | chronic |
| Q03 | congenital anomalies of neurological or sensory systems | chronic |
| Q04 | congenital anomalies of neurological or sensory systems | chronic |
| Q05 | congenital anomalies of neurological or sensory systems | chronic |
| Q06 | congenital anomalies of neurological or sensory systems | chronic |
| Q07 | congenital anomalies of neurological or sensory systems | chronic |
| Q10.4 | congenital anomalies of neurological or sensory systems | chronic |
| Q10.7 | congenital anomalies of neurological or sensory systems | chronic |
| Q11 | congenital anomalies of neurological or sensory systems | chronic |
| Q12 | congenital anomalies of neurological or sensory systems | chronic |
| Q13.0 | congenital anomalies of neurological or sensory systems | chronic |
| Q13.1 | congenital anomalies of neurological or sensory systems | chronic |
| Q13.2 | congenital anomalies of neurological or sensory systems | chronic |
| Q13.3 | congenital anomalies of neurological or sensory systems | chronic |
| Q13.4 | congenital anomalies of neurological or sensory systems | chronic |
| Q13.8 | congenital anomalies of neurological or sensory systems | chronic |
| Q13.9 | congenital anomalies of neurological or sensory systems | chronic |
| Q14 | congenital anomalies of neurological or sensory systems | chronic |
| Q15 | congenital anomalies of neurological or sensory systems | chronic |
| Q16 | congenital anomalies of neurological or sensory systems | chronic |
| Q75.0 | congenital anomalies of neurological or sensory systems | chronic |
| Q75.1 | congenital anomalies of neurological or sensory systems | chronic |
| Q85 | congenital anomalies of neurological or sensory systems | chronic |
| Q86.0 | congenital anomalies of neurological or sensory systems | chronic |
| Q86.1 | congenital anomalies of neurological or sensory systems | chronic |
| Q86.8 | congenital anomalies of neurological or sensory systems | chronic |
| Q90 | congenital anomalies of neurological or sensory systems | chronic |
| Q91 | congenital anomalies of neurological or sensory systems | chronic |
| Q92 | congenital anomalies of neurological or sensory systems | chronic |
| Q93 | congenital anomalies of neurological or sensory systems | chronic |
| Q95.2 | congenital anomalies of neurological or sensory systems | chronic |
| Q95.3 | congenital anomalies of neurological or sensory systems | chronic |
| Q97 | congenital anomalies of neurological or sensory systems | chronic |
| Q99 | congenital anomalies of neurological or sensory systems | chronic |
| F02.2 | other [neurological] | chronic |
| F02.3 | other [neurological] | chronic |
| G00 | other [neurological] | chronic |
| G01 | other [neurological] | chronic |
| G02 | other [neurological] | chronic |
| G03 | other [neurological] | chronic |
| G04 | other [neurological] | chronic |
| G05 | other [neurological] | chronic |
| G06 | other [neurological] | chronic |
| G07 | other [neurological] | chronic |
| G08 | other [neurological] | chronic |
| G09 | other [neurological] | chronic |
| G10 | other [neurological] | chronic |
| G11 | other [neurological] | chronic |
| G12 | other [neurological] | chronic |
| G13.8 | other [neurological] | chronic |
| G14 | other [neurological] | chronic |
| G20 | other [neurological] | chronic |
| G21 | other [neurological] | chronic |
| G22 | other [neurological] | chronic |
| G23 | other [neurological] | chronic |
| G24.1 | other [neurological] | chronic |
| G24.2 | other [neurological] | chronic |
| G24.3 | other [neurological] | chronic |
| G24.4 | other [neurological] | chronic |
| G24.5 | other [neurological] | chronic |
| G24.8 | other [neurological] | chronic |
| G24.9 | other [neurological] | chronic |
| G25 | other [neurological] | chronic |
| G26 | other [neurological] | chronic |
| G30 | other [neurological] | chronic |
| G31.0 | other [neurological] | chronic |
| G31.1 | other [neurological] | chronic |
| G31.8 | other [neurological] | chronic |
| G31.9 | other [neurological] | chronic |
| G32 | other [neurological] | chronic |
| G35 | other [neurological] | chronic |
| G36 | other [neurological] | chronic |
| G37 | other [neurological] | chronic |
| G45 | other [neurological] | chronic |
| G46 | other [neurological] | chronic |
| G50 | other [neurological] | chronic |
| G51 | other [neurological] | chronic |
| G52 | other [neurological] | chronic |
| G53.0 | other [neurological] | chronic |
| G53.1 | other [neurological] | chronic |
| G53.8 | other [neurological] | chronic |
| G54 | other [neurological] | chronic |
| G55.8 | other [neurological] | chronic |
| G56 | other [neurological] | chronic |
| G57 | other [neurological] | chronic |
| G58 | other [neurological] | chronic |
| G59.8 | other [neurological] | chronic |
| G60 | other [neurological] | chronic |
| G61 | other [neurological] | chronic |
| G62.0 | other [neurological] | chronic |
| G62.2 | other [neurological] | chronic |
| G62.8 | other [neurological] | chronic |
| G62.9 | other [neurological] | chronic |
| G64 | other [neurological] | chronic |
| G70 | other [neurological] | chronic |
| G71 | other [neurological] | chronic |
| G72.2 | other [neurological] | chronic |
| G72.3 | other [neurological] | chronic |
| G72.4 | other [neurological] | chronic |
| G72.8 | other [neurological] | chronic |
| G72.9 | other [neurological] | chronic |
| G73.0 | other [neurological] | chronic |
| G73.3 | other [neurological] | chronic |
| G90 | other [neurological] | chronic |
| G91 | other [neurological] | chronic |
| G92 | other [neurological] | chronic |
| G93 | other [neurological] | chronic |
| G94.2 | other [neurological] | chronic |
| G94.8 | other [neurological] | chronic |
| G95 | other [neurological] | chronic |
| G96 | other [neurological] | chronic |
| G98 | other [neurological] | chronic |
| G99.1 | other [neurological] | chronic |
| G99.2 | other [neurological] | chronic |
| I60 | other [neurological] | chronic |
| I61 | other [neurological] | chronic |
| I62 | other [neurological] | chronic |
| I63 | other [neurological] | chronic |
| I64 | other [neurological] | chronic |
| I65 | other [neurological] | chronic |
| I66 | other [neurological] | chronic |
| I67 | other [neurological] | chronic |
| I68.0 | other [neurological] | chronic |
| I68.2 | other [neurological] | chronic |
| I69 | other [neurological] | chronic |
| I72.0 | other [neurological] | chronic |
| I72.5 | other [neurological] | chronic |
| T85.0 | other [neurological] | chronic |
| T85.1 | other [neurological] | chronic |
| Y46.7 | other [neurological] | chronic |
| Y46.8 | other [neurological] | chronic |
| Z98.2 | other [neurological] | chronic |
| Q20 | congenital heart disease | chronic |
| Q21 | congenital heart disease | chronic |
| Q22 | congenital heart disease | chronic |
| Q23 | congenital heart disease | chronic |
| Q24 | congenital heart disease | chronic |
| Q25 | congenital heart disease | chronic |
| Q26 | congenital heart disease | chronic |
| Q89.3 | congenital heart disease | chronic |
| I00 | other [cardiovascular] | chronic |
| I01 | other [cardiovascular] | chronic |
| I02 | other [cardiovascular] | chronic |
| I05 | other [cardiovascular] | chronic |
| I06 | other [cardiovascular] | chronic |
| I07 | other [cardiovascular] | chronic |
| I08 | other [cardiovascular] | chronic |
| I09 | other [cardiovascular] | chronic |
| I10 | other [cardiovascular] | chronic |
| I11 | other [cardiovascular] | chronic |
| I12 | other [cardiovascular] | chronic |
| I13 | other [cardiovascular] | chronic |
| I15 | other [cardiovascular] | chronic |
| I20 | other [cardiovascular] | chronic |
| I21 | other [cardiovascular] | chronic |
| I22 | other [cardiovascular] | chronic |
| I23 | other [cardiovascular] | chronic |
| I24 | other [cardiovascular] | chronic |
| I25 | other [cardiovascular] | chronic |
| I26 | other [cardiovascular] | chronic |
| I27 | other [cardiovascular] | chronic |
| I28 | other [cardiovascular] | chronic |
| I31 | other [cardiovascular] | chronic |
| I32 | other [cardiovascular] | chronic |
| I33 | other [cardiovascular] | chronic |
| I34 | other [cardiovascular] | chronic |
| I35 | other [cardiovascular] | chronic |
| I36 | other [cardiovascular] | chronic |
| I37 | other [cardiovascular] | chronic |
| I38 | other [cardiovascular] | chronic |
| I39 | other [cardiovascular] | chronic |
| I41 | other [cardiovascular] | chronic |
| I42.0 | other [cardiovascular] | chronic |
| I42.1 | other [cardiovascular] | chronic |
| I42.2 | other [cardiovascular] | chronic |
| I42.3 | other [cardiovascular] | chronic |
| I42.4 | other [cardiovascular] | chronic |
| I42.5 | other [cardiovascular] | chronic |
| I42.7 | other [cardiovascular] | chronic |
| I42.8 | other [cardiovascular] | chronic |
| I42.9 | other [cardiovascular] | chronic |
| I43.0 | other [cardiovascular] | chronic |
| I43.1 | other [cardiovascular] | chronic |
| I43.2 | other [cardiovascular] | chronic |
| I43.8 | other [cardiovascular] | chronic |
| I44.1 | other [cardiovascular] | chronic |
| I44.2 | other [cardiovascular] | chronic |
| I44.3 | other [cardiovascular] | chronic |
| I44.4 | other [cardiovascular] | chronic |
| I44.5 | other [cardiovascular] | chronic |
| I44.6 | other [cardiovascular] | chronic |
| I44.7 | other [cardiovascular] | chronic |
| I45.1 | other [cardiovascular] | chronic |
| I45.2 | other [cardiovascular] | chronic |
| I45.3 | other [cardiovascular] | chronic |
| I45.4 | other [cardiovascular] | chronic |
| I45.5 | other [cardiovascular] | chronic |
| I45.6 | other [cardiovascular] | chronic |
| I45.8 | other [cardiovascular] | chronic |
| I45.9 | other [cardiovascular] | chronic |
| I46 | other [cardiovascular] | chronic |
| I47 | other [cardiovascular] | chronic |
| I48 | other [cardiovascular] | chronic |
| I49 | other [cardiovascular] | chronic |
| I50 | other [cardiovascular] | chronic |
| I51 | other [cardiovascular] | chronic |
| I52.8 | other [cardiovascular] | chronic |
| I70 | other [cardiovascular] | chronic |
| I71 | other [cardiovascular] | chronic |
| I72.1 | other [cardiovascular] | chronic |
| I72.2 | other [cardiovascular] | chronic |
| I74.4 | other [cardiovascular] | chronic |
| I72.8 | other [cardiovascular] | chronic |
| I72.9 | other [cardiovascular] | chronic |
| I73 | other [cardiovascular] | chronic |
| I74 | other [cardiovascular] | chronic |
| I77 | other [cardiovascular] | chronic |
| I79.0 | other [cardiovascular] | chronic |
| I79.1 | other [cardiovascular] | chronic |
| I79.8 | other [cardiovascular] | chronic |
| I81 | other [cardiovascular] | chronic |
| I82 | other [cardiovascular] | chronic |
| I98 | other [cardiovascular] | chronic |
| I99 | other [cardiovascular] | chronic |
| M03.6 | other [cardiovascular] | chronic |
| N08.8 | other [cardiovascular] | chronic |
| Q27 | other [cardiovascular] | chronic |
| Q28 | other [cardiovascular] | chronic |
| S26 | other [cardiovascular] | chronic |
| T82.0 | other [cardiovascular] | chronic |
| T82.1 | other [cardiovascular] | chronic |
| T82.2 | other [cardiovascular] | chronic |
| T82.3 | other [cardiovascular] | chronic |
| T82.5 | other [cardiovascular] | chronic |
| T82.6 | other [cardiovascular] | chronic |
| T82.7 | other [cardiovascular] | chronic |
| T82.8 | other [cardiovascular] | chronic |
| T82.9 | other [cardiovascular] | chronic |
| T86.2 | other [cardiovascular] | chronic |
| Y60.5 | other [cardiovascular] | chronic |
| Y61.5 | other [cardiovascular] | chronic |
| Y62.5 | other [cardiovascular] | chronic |
| Y84.0 | other [cardiovascular] | chronic |
| Z45.0 | other [cardiovascular] | chronic |
| Z50.0 | other [cardiovascular] | chronic |
| Z94.1 | other [cardiovascular] | chronic |
| Z95 | other [cardiovascular] | chronic |
| R62 | codes indicating non-specific chronic condition | chronic |
| Z43.1 | codes indicating non-specific chronic condition | chronic |
| Z51.5 | codes indicating non-specific chronic condition | chronic |
| Z75.5 | codes indicating non-specific chronic condition | chronic |
| Z93.1 | codes indicating non-specific chronic condition | chronic |
| Z99.3 | codes indicating non-specific chronic condition | chronic |
