## Supplementary material for "Repeat Hospitalisation Following Admission for Mental Ill-health and Stress-Related Presentations in Children and Young People in England between 2014-2019: A Retrospective Cohort Study": S2

### Supporting information – Appendix S2

#### Section 1: Sample Flow

Supplementary Figure 1: Data retention and flow during data cleaning steps

**Initial dataset: all episodes and spells**

(Age 10-25, 1^st^ April 2013-21^st^ March 2019, duplicates removed, valid episode end date)

PIDs: 1,507,068

spells: 4,321,015

episodes: 5,158,597

**Index >24th birthday month**

PIDs: 121,991

spells: 331,980

episodes: 402,290

**Individuals without any MH/SRP spell**

(MH/SRP in non-prioritised diagnostic position or with medical exclusion)

PIDs: 776,878

spells: 2,051,702

episodes: 2,418,474

**Exclusions**

**Index later than 31^st^ March 2018**

PIDs: 78,015

spells: 184,984

episodes: 220,860

**Meeting diagnostic inclusion criteria**

PIDs: 730,190

spells: 2,269,313

episodes: 2,740,123

**Pre-index spells and non-index spells**

**> 365 days from index**

PIDs: 0

spells: 856,786

episodes: 1,037,417

**Index <24^th^ birthday month**

PIDs: 608,199

spells: 1,937,333

episodes: 2,337,833

**Index visit before 31st March 2018**

PIDs: 540,611

spells: 1,781,758

episodes: 2,149,632

**Deceased during index admission**

PIDs: 168

spells: 180

episodes314

**Index visits, and re-hospitalisations within 365 days from index**

PIDs: 540,611

spells: 924,972

episodes: 1,112,215

**Included data**

(MH/SRP spells, first presentation or 365 days from prior discharge, index up to 24 birthday month)

PIDs: 492,061

Total index visits: 518,504

Total included spells (index + readmissions within 365 days): 924,792

episodes: 1,111,901

#### Section 2: Missing data

##### Supplementary Table ST1: Missing data in key outcome measures and covariates on index admission

**Note:** Missingness is not present in some key outcome measures due to their definitions. Study inclusion criteria specify required age ranges, and index admissions are defined by specific diagnostic characteristics, excluding cases with missing values for these measures. Similarly chronic conditions are determined solely by the presence of relevant diagnostic codes at and prior to index presentations.

Missing data for discharge method includes both spells recorded as “Unfinished hospital spell” and those with missing entries.

| **Measure** | **Denominator** | **Datapoints missing (N)** | **Percentage missing (%)** |
| --- | --- | --- | --- |
| Sex | Number of patients | 0 | 0 |
| IMD | Number of patients | 9,171 | 1.51 |
| Region | Number of patients | 0 | 0 |
| Ethnicity | Number of patients | 60,752 | 10.01 |
| Age | Number of patients | 0 | 0 |
| Diagnosis group | Number of index spells | 0 | 0 |
| Chronic condition at index | Number of index spells | 0 | 0 |
| Admission method | Number of index spells | 619 | 0.1 |
| Discharge method | Number of index spells | 948 | 0.15 |
| Admission duration | Number of index spells | 0 | 0 |

#### Section 3: Chronic Conditions

##### Supplementary Table ST2: number of index presentations with co-occurring history or presence of chronic medical conditions

Chronic conditions identified via code list developed by Hardelid et al. (2014), after exclusion of mental health conditions. More detailed information provided in methods.

| **Diagnostic group at index** | **Number of index presentations** | **Number with history of chronic conditions** | **% Chronic history at index** |
| --- | --- | --- | --- |
| Eating disorders | 3,357 | 837 | 24.93 |
| Externalising | 560 | 155 | 27.68 |
| Internalising | 8,472 | 2,371 | 27.99 |
| Personality disorders | 1,019 | 262 | 25.71 |
| Postpartum mental health | 75 | 29 | 38.67 |
| Potentially psychosomatic | 351,759 | 137,613 | 39.12 |
| Psychosis | 6,905 | 1,190 | 17.23 |
| Self-harm | 131,653 | 36,943 | 28.06 |
| Substance use or abuse | 14,704 | 2,907 | 19.77 |

#### Section 4: Diagnostic stability and change during re-hospitalisation

##### Supplementary table ST3: Diagnostic stability and percentage of patients with MH/SRP or non-MH/SRP diagnosis on re-hospitalisation

Proportion of re-hospitalisations within the same diagnostic group as at first index presentation, proportion of subsequent presentations associated with MH/SRP presentations, and proportion of subsequent presentations with non-MH/SRP diagnostic codes for all outcomes. Excepting 1 year MH/SRP re-hospitalisations, where only readmissions for index group presentations only are shown (proportion with MH/SRP presentations are 100%, and proportion of non-MH/SRP presentations are 0% by definition). -† count suppressed to avoid disclosure.

| **Outcome** | **index diagnosis group** | **Re-Hospitalisation Diagnosis group** | | |
| --- | --- | --- | --- | --- |
|  |  | % Same group as index | % MH or SRP | % non-MH/SRP |
| 30 day all-cause re-hospitalisations | Eating disorders | 64.75 | 81.64 | 18.36 |
|  | Externalising | 24.44 | 71.11 | 28.89 |
|  | Internalising | 21.61 | 67.71 | 32.29 |
|  | Personality disorders | 31.40 | 84.88 | 15.12 |
|  | Postpartum mental health | -† | -† | -† |
|  | Potentially psychosomatic | 39.93 | 43.64 | 56.36 |
|  | Psychosis | 56.23 | 79.09 | 20.91 |
|  | Self-harm | 70.60 | 79.15 | 20.85 |
|  | Substance use/abuse | 25.37 | 63.59 | 36.41 |
|  | Overall | 47.25 | 53.76 | 46.24 |
| 1 year all-cause re-hospitalisations | Eating disorders | 52.72 | 77.49 | 22.51 |
|  | Externalising | 12.35 | 59.41 | 40.59 |
|  | Internalising | 12.83 | 54.90 | 45.10 |
|  | Personality disorders | 21.37 | 75.38 | 24.62 |
|  | Postpartum mental health | -† | -† | -† |
|  | Potentially psychosomatic | 30.03 | 34.86 | 65.14 |
|  | Psychosis | 47.62 | 72.18 | 27.82 |
|  | Self-harm | 59.41 | 69.60 | 30.40 |
|  | Substance use/abuse | 15.26 | 49.75 | 50.25 |
|  | Overall | 37.68 | 45.63 | 54.37 |
| 1 year MH/SRP only re-hospitalisations | Eating disorders | 66.67 | - | - |
|  | Externalising | 20.37 | - | - |
|  | Internalising | 22.54 |  |  |
|  | Personality disorders | 27.38 |  |  |
|  | Postpartum mental health | 22.22 |  |  |
|  | Potentially psychosomatic | 86.37 | - | - |
|  | Psychosis | 65.20 | - | - |
|  | Self-harm | 84.31 | - | - |
|  | Substance use/abuse | 30.09 | - | - |
|  | Overall | 82.45 | - | - |

##### Supplementary Figure SF2: alluvial plot for transition and stability of derived diagnostic groupings on successive admissions from index to all-cause hospitalisations within 30 days, with minimum transition frequency of n≥100.

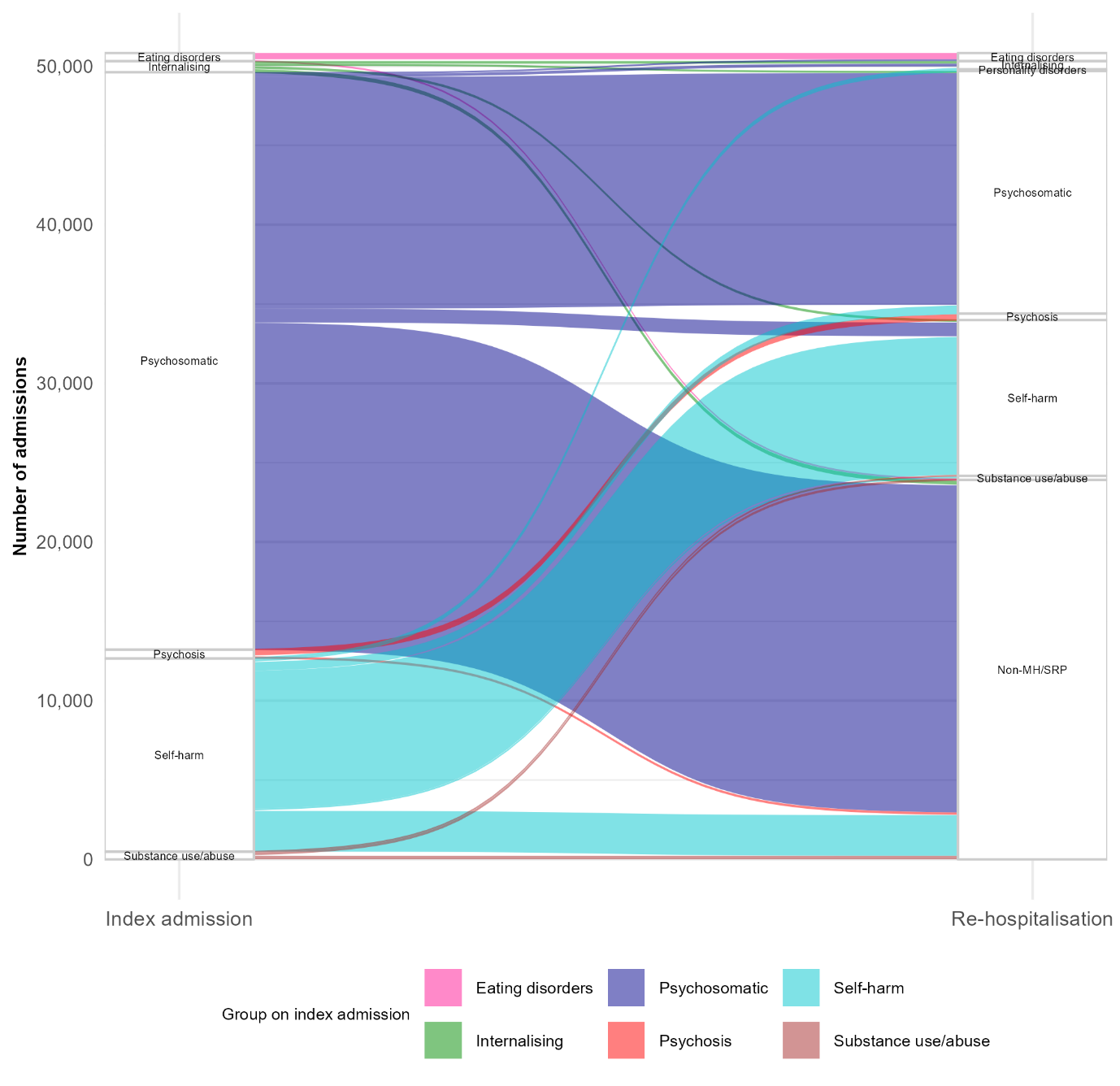

##### Supplementary Figure SF3: alluvial plot for transition and stability of derived diagnostic groupings on successive admissions from index to all-cause hospitalisations extracting MH/SRP presentations only, with minimum transition frequency of n≥100.

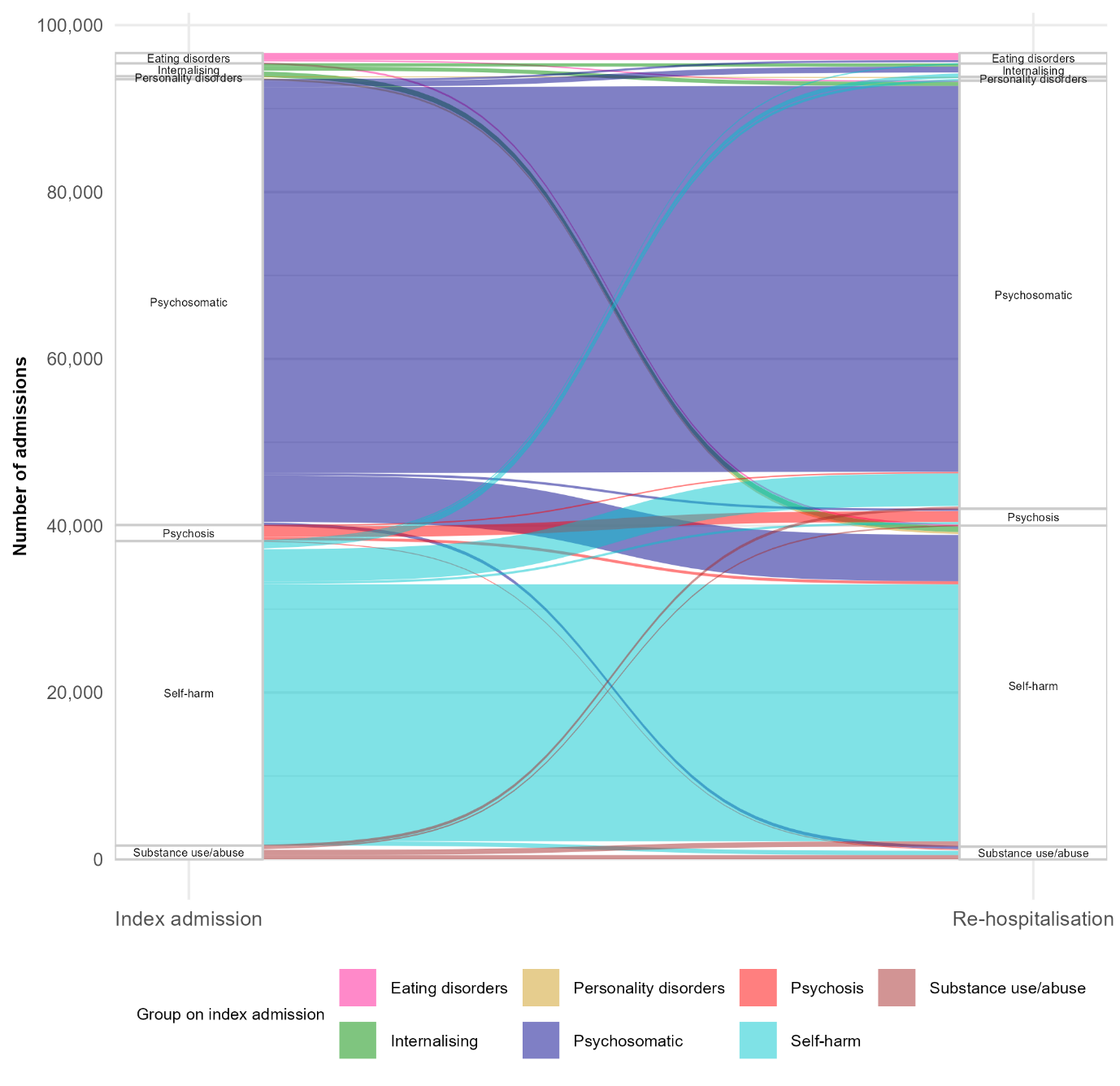

#### Section 5: Survival analysis

##### Supplementary Figure SF3: Sensitivity analysis: Cumulative monthly probability of all-cause readmission separated out by diagnostic group with tabulation below showing 1-month, 3-month, 6-month and 12-month readmission probability, together with 95% confidence intervals, excluding repeat index admissions

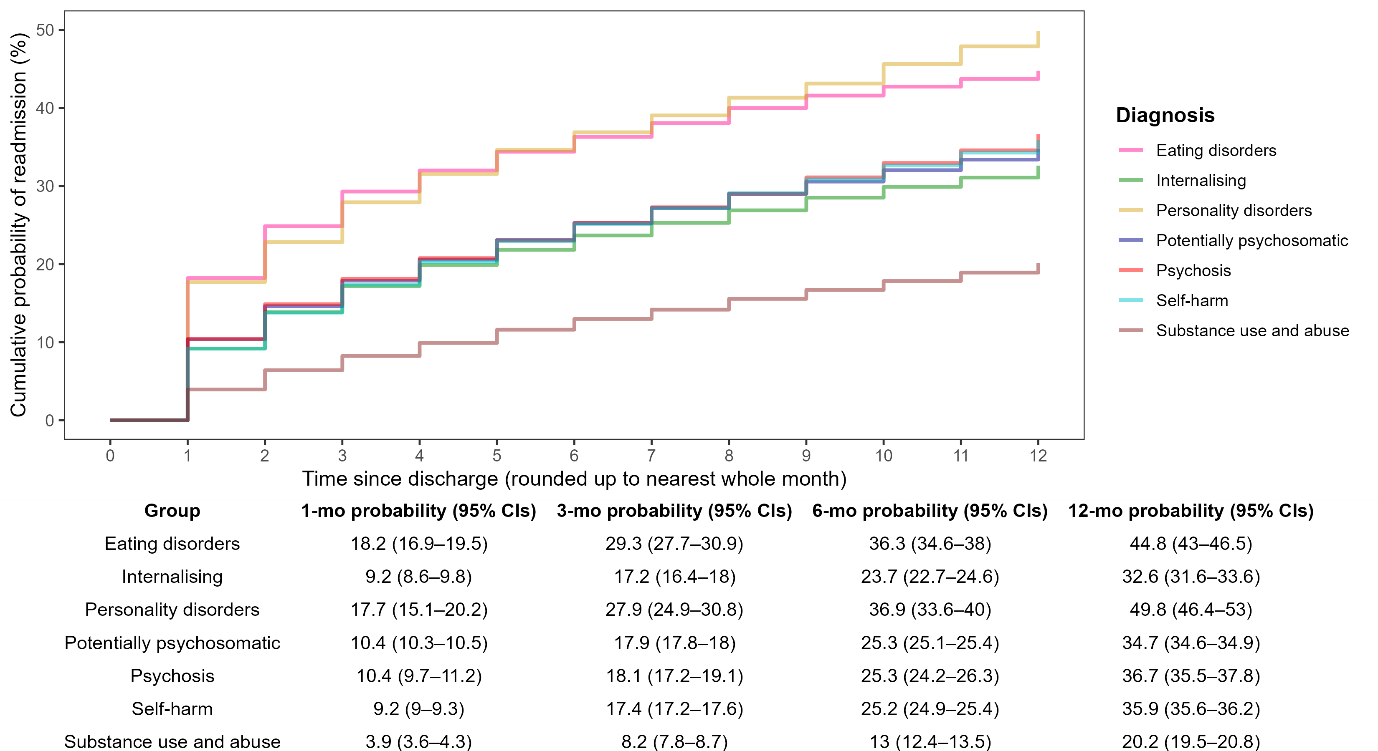

#### Section 6: Regression results

##### Supplementary Table ST4: GEE results predicting rehospitalisation: complete case analysis

Odds ratios (OR) for minimally adjusted model (Min adj.) and ORs for fully adjusted models with 95% confidence intervals and p-values for regression predictors.

| **Measure:** reference group | **Subgroup** | **All-cause readmissions** | | | | | | | | **MH/SRP only readmissions** | | | |
| --- | --- | --- | --- | --- | --- | --- | --- | --- | --- | --- | --- | --- | --- |
|  |  | **Readmission within 30 days** | | | | **Readmission within 1 year** | | | | **Readmission within 1 year** | | | |
|  |  | Min adj. OR | Fully adjusted model | | | Min adj. OR | Fully adjusted model | | | Min adj. OR | Fully adjusted model | | |
|  |  |  | OR | 95% CI | p value |  | OR | 95% CI | p value |  | OR | 95% CI | p value |
| **Sex:** Male | Female | 1.23 | 1.19 | 1.16–1.21 | <0.001 | 1.64 | 1.57 | 1.55–1.6 | <0.001 | 1.52 | 1.39 | 1.37–1.42 | <0.001 |
| **Age at index:** 10-12 | 12-13 | 0.99 | 0.98 | 0.94–1.03 | 0.41 | 1.17 | 1.11 | 1.07–1.14 | <0.001 | 1.48 | 1.21 | 1.16–1.26 | <0.001 |
|  | 14-15 | 0.95 | 0.95 | 0.91–0.99 | 0.02 | 1.31 | 1.18 | 1.15–1.22 | <0.001 | 1.90 | 1.3 | 1.25–1.35 | <0.001 |
|  | 16-17 | 0.90 | 0.86 | 0.82–0.9 | <0.001 | 1.35 | 1.16 | 1.13–1.2 | <0.001 | 1.60 | 1.09 | 1.05–1.13 | <0.001 |
|  | 18-19 | 0.97 | 0.9 | 0.86–0.94 | <0.001 | 1.53 | 1.27 | 1.24–1.31 | <0.001 | 1.56 | 1.08 | 1.04–1.12 | <0.001 |
|  | 20-21 | 1.00 | 0.9 | 0.86–0.93 | <0.001 | 1.65 | 1.32 | 1.29–1.36 | <0.001 | 1.46 | 1.02 | 0.98–1.06 | 0.26 |
|  | 22-24 | 1.02 | 0.89 | 0.85–0.93 | <0.001 | 1.75 | 1.37 | 1.33–1.41 | <0.001 | 1.44 | 1.01 | 0.98–1.05 | 0.55 |
| **Ethnicity:** White | Asian | 1.1 | 1.07 | 1.03–1.11 | <0.001 | 0.88 | 0.87 | 0.85–0.89 | <0.001 | 0.80 | 0.86 | 0.83–0.88 | <0.001 |
|  | Black | 1.08 | 1.04 | 0.99–1.1 | 0.11 | 1.00 | 0.94 | 0.91–0.98 | 0.00 | 0.91 | 0.89 | 0.86–0.93 | <0.001 |
|  | Mixed | 0.98 | 0.97 | 0.91–1.03 | 0.30 | 0.98 | 0.96 | 0.92–1 | 0.07 | 1.01 | 0.95 | 0.91–1.00 | 0.05 |
|  | Other | 0.88 | 0.94 | 0.88–1 | 0.05 | 0.75 | 0.79 | 0.76–0.82 | <0.001 | 0.73 | 0.78 | 0.74–0.82 | <0.001 |
| **IMD:**  5 = least deprived | 1 = most deprived | 1.11 | 1.07 | 1.03–1.1 | <0.001 | 1.38 | 1.35 | 1.32–1.38 | <0.001 | 1.19 | 1.17 | 1.14–1.20 | <0.001 |
|  | 2 | 1.04 | 1.01 | 0.98–1.04 | 0.56 | 1.23 | 1.2 | 1.17–1.22 | <0.001 | 1.10 | 1.08 | 1.05–1.11 | <0.001 |
|  | 3 | 1.04 | 1.02 | 0.98–1.05 | 0.37 | 1.15 | 1.13 | 1.11–1.16 | <0.001 | 1.08 | 1.06 | 1.03–1.08 | <0.001 |
|  | 4 | 1.02 | 1.01 | 0.97–1.04 | 0.73 | 1.08 | 1.07 | 1.05–1.1 | <0.001 | 1.03 | 1.02 | 0.99–1.05 | 0.15 |
| **Region:**  South East | East Midlands | 1.02 | 0.99 | 0.95–1.03 | 0.67 | 1.13 | 1.05 | 1.02–1.09 | <0.001 | 1.07 | 1.01 | 0.98–1.04 | 0.57 |
|  | East of England | 1.04 | 1.01 | 0.97–1.05 | 0.50 | 1.06 | 1.03 | 1–1.05 | 0.05 | 1.01 | 1 | 0.97–1.03 | 0.88 |
|  | London | 1.05 | 1.01 | 0.97–1.05 | 0.60 | 1.04 | 1.01 | 0.98–1.04 | 0.44 | 0.94 | 0.97 | 0.94–1.00 | 0.08 |
|  | North East | 0.97 | 0.94 | 0.9–0.99 | 0.02 | 1.06 | 0.98 | 0.95–1.01 | 0.25 | 0.93 | 0.88 | 0.85–0.92 | <0.001 |
|  | North West | 1.08 | 1.03 | 0.99–1.07 | 0.12 | 1.13 | 1.02 | 1–1.04 | 0.11 | 1.05 | 0.98 | 0.95–1.01 | 0.13 |
|  | South West | 1.07 | 1.05 | 1.01–1.09 | 0.02 | 1.09 | 1.03 | 1.01–1.06 | 0.01 | 1.14 | 1.07 | 1.04–1.10 | <0.001 |
|  | West Midlands | 1.02 | 0.99 | 0.95–1.02 | 0.44 | 1 | 0.94 | 0.91–0.96 | <0.001 | 0.97 | 0.95 | 0.92–0.98 | <0.001 |
|  | Yorkshire and Humber | 1.10 | 1.05 | 1.01–1.09 | 0.02 | 1.09 | 0.99 | 0.96–1.01 | 0.29 | 0.94 | 0.91 | 0.88–0.93 | <0.001 |
| **Diagnosis:** Potentially psychosomatic | Eating disorders | 1.95 | 2.25 | 2.05–2.48 | <0.001 | 1.52 | 1.89 | 1.75–2.05 | <0.001 | 3.39 | 3.55 | 3.28–3.83 | <0.001 |
|  | Externalising | 0.78 | 0.92 | 0.67–1.27 | 0.61 | 0.82 | 1.19 | 0.98–1.46 | 0.09 | 1.32 | 1.61 | 1.29–2.03 | <0.001 |
|  | Internalising | 0.86 | 0.97 | 0.9–1.06 | 0.52 | 0.91 | 1.09 | 1.03–1.14 | 0.00 | 1.46 | 1.64 | 1.55–1.74 | <0.001 |
|  | Personality disorders | 1.73 | 2.1 | 1.76–2.5 | <0.001 | 1.84 | 2.2 | 1.92–2.53 | <0.001 | 3.97 | 4.9 | 4.29–5.60 | <0.001 |
|  | Psychosis | 0.97 | 1.33 | 1.22–1.45 | <0.001 | 1.08 | 1.65 | 1.56–1.75 | <0.001 | 2.21 | 3.29 | 3.10–3.50 | <0.001 |
|  | Self-harm | 0.89 | 0.99 | 0.96–1.01 | 0.24 | 1.06 | 1.18 | 1.16–1.2 | <0.001 | 2.14 | 2.23 | 2.2–2.27 | <0.001 |
|  | Substance use and abuse | 0.39 | 0.5 | 0.46–0.55 | <0.001 | 0.52 | 0.71 | 0.68–0.74 | <0.001 | 0.80 | 0.99 | 0.94–1.05 | 0.76 |
| **Chronic health condition**: No | Yes | 2.67 | 2.67 | 2.62–2.73 | <0.001 | 3.77 | 3.76 | 3.72–3.81 | <0.001 | 2.04 | 2.32 | 2.28–2.35 | <0.001 |
| **Admission method**: Non-emergency | Emergency | 0.96 | 1.1 | 1.07–1.13 | <0.001 | 0.82 | 0.97 | 0.95–0.98 | <0.001 | 1.10 | 1.1 | 1.07–1.12 | <0.001 |
| **Discharge method**:  formal discharge | Self-discharged | 1.06 | 1.08 | 1.02–1.14 | 0.01 | 1.10 | 1.05 | 1.01–1.09 | 0.01 | 1.09 | 1.04 | 1.00–1.09 | 0.07 |
| **Fiscal year of index presentation**:  2014-2015 | 2015-2016 | 0.89 | 0.89 | 0.87–0.92 | <0.001 | 0.86 | 0.86 | 0.84–0.87 | <0.001 | 0.84 | 0.84 | 0.82–0.86 | <0.001 |
|  | 2016-2017 | 0.86 | 0.85 | 0.83–0.88 | <0.001 | 0.83 | 0.82 | 0.8–0.83 | <0.001 | 0.81 | 0.82 | 0.80–0.83 | <0.001 |
|  | 2017-2018 | 0.88 | 0.87 | 0.84–0.89 | <0.001 | 0.8 | 0.78 | 0.77–0.8 | <0.001 | 0.83 | 0.82 | 0.81–0.84 | <0.001 |

##### Supplementary Table ST5: GEE results predicting rehospitalisation: sensitivity analysis including missing data groups

Odds ratios (OR) for minimally adjusted model (Min adj.) and ORs for fully adjusted models with 95% confidence intervals and p-values for regression predictors, including missing data categories.

| **Measure:** reference group | **Subgroup** | **All-cause readmissions** | | | | | | | | **MH/SRP only readmissions** | | | |
| --- | --- | --- | --- | --- | --- | --- | --- | --- | --- | --- | --- | --- | --- |
|  |  | **Readmission within 30 days** | | | | **Readmission within 1 year** | | | | **Readmission within 1 year** | | | |
|  |  | Min adj. OR | Fully adjusted model | | | Min adj. OR | Fully adjusted model | | | Min adj. OR | Fully adjusted model | | |
|  |  |  | OR | 95% CI | p value |  | OR | 95% CI | p value |  | OR | 95% CI | p value |
| **Sex:** Male | Female | 1.24 | 1.19 | 1.16–1.21 | <0.001 | 1.65 | 1.57 | 1.55–1.59 | <0.001 | 1.54 | 1.4 | 1.38–1.42 | <0.001 |
| **Age at index:** 10-12 | 12-13 | 0.99 | 0.98 | 0.93–1.02 | 0.33 | 1.16 | 1.1 | 1.07–1.14 | <0.001 | 1.47 | 1.21 | 1.16–1.25 | <0.001 |
|  | 14-15 | 0.94 | 0.94 | 0.9–0.99 | 0.01 | 1.30 | 1.18 | 1.15–1.22 | <0.001 | 1.88 | 1.3 | 1.25–1.35 | <0.001 |
|  | 16-17 | 0.89 | 0.86 | 0.82–0.9 | <0.001 | 1.32 | 1.16 | 1.12–1.19 | <0.001 | 1.57 | 1.09 | 1.05–1.13 | <0.001 |
|  | 18-19 | 0.95 | 0.89 | 0.85–0.93 | <0.001 | 1.46 | 1.25 | 1.22–1.29 | <0.001 | 1.51 | 1.07 | 1.03–1.1 | <0.001 |
|  | 20-21 | 0.98 | 0.89 | 0.85–0.93 | <0.001 | 1.56 | 1.3 | 1.26–1.33 | <0.001 | 1.41 | 1.01 | 0.97–1.04 | 0.77 |
|  | 22-24 | 0.99 | 0.88 | 0.85–0.92 | <0.001 | 1.66 | 1.34 | 1.3–1.38 | <0.001 | 1.39 | 1 | 0.96–1.03 | 0.80 |
| **Ethnicity:** White | Asian | 1.10 | 1.06 | 1.02–1.11 | 0.00 | 0.88 | 0.87 | 0.85–0.89 | <0.001 | 0.80 | 0.85 | 0.83–0.88 | <0.001 |
|  | Black | 1.08 | 1.05 | 0.99–1.1 | 0.08 | 1.00 | 0.95 | 0.92–0.98 | 0.00 | 0.92 | 0.9 | 0.87–0.94 | <0.001 |
|  | Mixed | 0.99 | 0.97 | 0.91–1.03 | 0.29 | 0.98 | 0.96 | 0.92–1 | 0.06 | 1.01 | 0.95 | 0.91–1 | 0.05 |
|  | Other | 0.86 | 0.93 | 0.87–0.99 | 0.03 | 0.72 | 0.78 | 0.75–0.82 | <0.001 | 0.71 | 0.77 | 0.74–0.81 | <0.001 |
|  | Missing | 0.46 | 0.54 | 0.52–0.57 | <0.001 | 0.34 | 0.39 | 0.38–0.4 | <0.001 | 0.40 | 0.46 | 0.45–0.47 | <0.001 |
| **IMD:**  5 = least deprived | 1 = most deprived | 1.13 | 1.07 | 1.04–1.1 | <0.001 | 1.4 | 1.35 | 1.32–1.38 | <0.001 | 1.21 | 1.17 | 1.14–1.2 | <0.001 |
|  | 2 | 1.05 | 1.01 | 0.98–1.05 | 0.37 | 1.24 | 1.2 | 1.18–1.23 | <0.001 | 1.11 | 1.08 | 1.05–1.11 | <0.001 |
|  | 3 | 1.04 | 1.02 | 0.99–1.05 | 0.26 | 1.16 | 1.14 | 1.11–1.16 | <0.001 | 1.08 | 1.06 | 1.03–1.08 | <0.001 |
|  | 4 | 1.02 | 1.01 | 0.98–1.04 | 0.59 | 1.09 | 1.08 | 1.05–1.1 | <0.001 | 1.03 | 1.02 | 0.99–1.05 | 0.11 |
|  | Missing | 0.40 | 0.49 | 0.43–0.55 | <0.001 | 0.29 | 0.35 | 0.32–0.37 | <0.001 | 0.32 | 0.38 | 0.35–0.42 | <0.001 |
| **Region:**  South East | East Midlands | 1.03 | 0.99 | 0.95–1.03 | 0.52 | 1.15 | 1.05 | 1.02–1.08 | <0.001 | 1.07 | 1 | 0.97–1.03 | 0.97 |
|  | East of England | 1.05 | 1.01 | 0.97–1.05 | 0.67 | 1.07 | 1.02 | 0.99–1.05 | 0.12 | 1.02 | 1 | 0.97–1.03 | 0.99 |
|  | London | 1.04 | 1 | 0.96–1.04 | 0.99 | 1.02 | 0.99 | 0.97–1.02 | 0.47 | 0.93 | 0.97 | 0.94–0.99 | 0.02 |
|  | North East | 0.99 | 0.94 | 0.9–0.99 | 0.01 | 1.10 | 0.97 | 0.94–1.01 | 0.10 | 0.97 | 0.88 | 0.85–0.92 | <0.001 |
|  | North West | 1.09 | 1.03 | 0.99–1.06 | 0.10 | 1.14 | 1.01 | 0.99–1.04 | 0.29 | 1.06 | 0.98 | 0.95–1 | 0.07 |
|  | South West | 1.07 | 1.05 | 1.01–1.09 | 0.01 | 1.07 | 1.03 | 1–1.05 | 0.04 | 1.13 | 1.07 | 1.04–1.1 | <0.001 |
|  | West Midlands | 1.04 | 0.99 | 0.95–1.03 | 0.57 | 1.02 | 0.93 | 0.91–0.96 | <0.001 | 0.98 | 0.95 | 0.92–0.98 | <0.001 |
|  | Yorkshire and Humber | 1.10 | 1.04 | 1–1.08 | 0.07 | 1.09 | 0.98 | 0.95–1 | 0.07 | 0.95 | 0.91 | 0.88–0.93 | <0.001 |
| **Diagnosis:** Potentially psychosomatic | Eating disorders | 1.91 | 2.18 | 1.99–2.39 | <0.001 | 1.50 | 1.83 | 1.7–1.98 | <0.001 | 3.32 | 3.42 | 3.18–3.69 | <0.001 |
|  | Externalising | 0.75 | 0.87 | 0.64–1.18 | 0.37 | 0.81 | 1.12 | 0.93–1.36 | 0.24 | 1.34 | 1.59 | 1.28–1.98 | <0.001 |
|  | Internalising | 0.86 | 0.98 | 0.91–1.06 | 0.63 | 0.91 | 1.08 | 1.03–1.13 | 0.00 | 1.43 | 1.62 | 1.53–1.71 | <0.001 |
|  | Personality disorder | 1.71 | 2.03 | 1.71–2.41 | <0.001 | 1.90 | 2.24 | 1.97–2.55 | <0.001 | 3.94 | 4.77 | 4.2–5.41 | <0.001 |
|  | Psychosis | 1.00 | 1.35 | 1.25–1.47 | <0.001 | 1.09 | 1.65 | 1.56–1.74 | <0.001 | 2.22 | 3.28 | 3.09–3.47 | <0.001 |
|  | Self-harm | 0.89 | 0.98 | 0.95–1 | 0.05 | 1.06 | 1.18 | 1.16–1.2 | <0.001 | 2.13 | 2.22 | 2.19–2.26 | <0.001 |
|  | Substance use and abuse | 0.37 | 0.49 | 0.45–0.53 | <0.001 | 0.48 | 0.7 | 0.67–0.73 | <0.001 | 0.73 | 0.97 | 0.92–1.02 | 0.25 |
| **Chronic health condition**: No | Yes | 2.75 | 2.68 | 2.63–2.73 | <0.001 | 3.87 | 3.75 | 3.7–3.8 | <0.001 | 2.11 | 2.31 | 2.27–2.34 | <0.001 |
| **Admission method**: Non-emergency | Emergency | 0.97 | 1.09 | 1.06–1.12 | <0.001 | 0.83 | 0.94 | 0.93–0.96 | <0.001 | 1.09 | 1.06 | 1.04–1.09 | <0.001 |
|  | Missing | 0.89 | 0.95 | 0.71–1.28 | 0.76 | 0.95 | 1.06 | 0.89–1.27 | 0.53 | 1.64 | 1.09 | 0.9–1.33 | 0.37 |
| **Discharge method**:  formal discharge | Self-discharged | 1.05 | 1.07 | 1.01–1.13 | 0.02 | 1.08 | 1.05 | 1.01–1.09 | 0.02 | 1.09 | 1.05 | 1–1.1 | 0.03 |
|  | Missing | 1.18 | 1.42 | 1.19–1.69 | <0.001 | 1.10 | 1.35 | 1.2–1.52 | <0.001 | 1.52 | 1.43 | 1.26–1.63 | <0.001 |
| **Fiscal year of index presentation**:  2014-2015 | 2015-2016 | 0.89 | 0.9 | 0.88–0.92 | <0.001 | 0.86 | 0.87 | 0.85–0.88 | <0.001 | 0.84 | 0.85 | 0.83–0.86 | <0.001 |
|  | 2016-2017 | 0.86 | 0.86 | 0.84–0.89 | <0.001 | 0.84 | 0.83 | 0.82–0.85 | <0.001 | 0.82 | 0.83 | 0.81–0.85 | <0.001 |
|  | 2017-2018 | 0.88 | 0.87 | 0.85–0.9 | <0.001 | 0.81 | 0.8 | 0.78–0.81 | <0.001 | 0.84 | 0.84 | 0.82–0.86 | <0.001 |

##### Supplementary Table ST6: Poisson regression results predicting frequency of all-cause readmission within 1 year of index admission

| **Predictor:** Reference group | **Subgroup** | **IRR** | **95% CIs** | **P-value** |
| --- | --- | --- | --- | --- |
| **Sex:** Male | Female | 1.05 | 1.04–1.06 | <0.001 |
| **Age at index:** 10-12 | 12-13 | 1.03 | 1.00–1.05 | 0.06 |
|  | 14-15 | 1.03 | 1.01–1.05 | 0.05 |
|  | 16-17 | 1 | 0.98–1.02 | 1 |
|  | 18-19 | 1 | 0.98–1.03 | 0.82 |
|  | 20-21 | 1 | 0.98–1.02 | 0.83 |
|  | 22-24 | 1 | 0.97–1.02 | 0.72 |
| **Ethnicity:** White | Asian | 0.97 | 0.95–0.99 | 0.02 |
|  | Black | 0.95 | 0.92–0.97 | <0.001 |
|  | Mixed | 0.95 | 0.92–0.98 | <0.01 |
|  | Other | 0.93 | 0.90–0.96 | <0.001 |
| **IMD:** 5 = least deprived | 1 = most deprived | 1 | 0.98–1.02 | 0.97 |
|  | 2 | 1 | 0.99–1.02 | 0.59 |
|  | 3 | 1.01 | 0.99–1.02 | 0.53 |
|  | 4 | 1 | 0.98–1.02 | 0.86 |
| **Region:** South East | East Midlands | 0.98 | 0.96–1.00 | 0.06 |
|  | East of England | 0.98 | 0.96–1.00 | 0.06 |
|  | London | 0.98 | 0.96–1.00 | 0.1 |
|  | North East | 0.98 | 0.96–1.00 | 0.11 |
|  | North West | 0.99 | 0.97–1.01 | 0.28 |
|  | South West | 1.04 | 1.02–1.06 | <0.001 |
|  | West Midlands | 0.99 | 0.97–1.01 | 0.32 |
|  | Yorkshire and Humber | 1 | 0.98–1.02 | 0.92 |
| **Diagnosis:** Potentially psychosomatic | Eating disorders | 1.21 | 1.16–1.26 | <0.001 |
|  | Externalising | 1.17 | 1.04–1.33 | 0.04 |
|  | Internalising | 1.15 | 1.12–1.20 | <0.001 |
|  | Personality disorders | 1.7 | 1.59–1.83 | <0.001 |
|  | Psychosis | 1.13 | 1.09–1.16 | <0.001 |
|  | Self-harm | 1.3 | 1.29–1.31 | <0.001 |
|  | Substance use and abuse | 1.14 | 1.10–1.18 | <0.001 |
| **Chronic health condition**: No | Yes | 1.2 | 1.19–1.21 | <0.001 |
| **Admission method**: Non-emergency | Emergency | 1.04 | 1.03–1.05 | <0.001 |
| **Discharge method**: formal discharge | Self-discharged | 1.01 | 0.98–1.04 | 0.43 |
| **Fiscal year of index presentation**: 2014-2015 | 2015-2016 | 0.91 | 0.90–0.92 | <0.001 |
|  | 2016-2017 | 0.91 | 0.89–0.92 | <0.001 |
|  | 2017-2018 | 0.91 | 0.90–0.92 | <0.001 |
